# Effectiveness of mRNA COVID-19 vaccine boosters against infection, hospitalization and death: a target trial emulation in the omicron (B.1.1.529) variant era

**DOI:** 10.1101/2022.06.15.22276466

**Authors:** George N. Ioannou, Amy SB Bohnert, Ann M. O’Hare, Edward J. Boyko, Matthew L. Maciejewski, Valerie A. Smith, C. Barrett Bowling, Elizabeth Viglianti, Theodore J. Iwashyna, Denise M. Hynes, Kristin Berry, the COVID-19 Observational Research Collaboratory (CORC)

## Abstract

**Abstract:** *Background:* The effectiveness of a 3^rd^ mRNA COVID-19 vaccine (“booster”) dose against the omicron (B.1.1.529) variant is uncertain especially in older, high-risk populations.

*Objective:* To determine mRNA booster vaccine effectiveness (VE) against SARS-CoV-2 infection, hospitalization and death in the omicron era by type of booster, type of primary vaccine, time since primary vaccine, age and comorbidity burden.

*Design:* Target trial emulation study comparing booster vaccination versus no booster.

*Setting:* U.S. Department of Veterans Affairs (VA) healthcare system

*Participants and Intervention:* Among persons who had received two mRNA COVID-19 vaccine doses at least 5 months earlier, we designed this retrospective matched cohort study to emulate a target trial of booster mRNA vaccination (BNT162b2 or mRNA-1273) versus no booster, conducted from 12/01/2021 to 03/31/2022.

*Measurements:* Booster VE.

*Results:* Each group included 490,838 well-matched persons, predominantly male (88%), mean age 63.0±14.0 years, followed for up to 121 days (mean 79.8 days). Booster VE >10 days after booster was 42.3% (95% CI 40.6-43.9) against SARS-CoV-2 infection, 53.3% (48.1-58.0) against SARS-CoV-2-related hospitalization and 79.1% (71.2-84.9) against SARS-CoV-2-related death. Booster VE was similar for different booster types (BNT162b2 or mRNA-1273), age groups or primary vaccination regimens, but was significantly higher with longer time since primary vaccination and with higher comorbidity burden.

*Limitations:* Predominantly male population.

*Conclusions:* Booster mRNA vaccination was highly effective in preventing death and moderately effective in preventing infection and hospitalization for up to 4 months after administration in the omicron era. Increased uptake of booster vaccination, which is currently suboptimal, should be pursued to limit the morbidity and mortality of SARS-CoV-2 infection, especially in persons with high comorbidity burden. **Primary Funding Source:** Department of Veterans Affairs

## Introduction

The Centers for Disease Control and Prevention (CDC) recommends that all persons 5 years and older receive a 3^rd^ dose (“booster”) of an mRNA COVID-19 vaccine (Pfizer-BioNTech’s BNT162b2 or Moderna’s mRNA-1273) at least 5 months after receipt of the second mRNA vaccine dose^1^. Despite these recommendations, uptake of booster vaccination in the United States remains low at ∼35.8% as of 6/12/2022^2^.

A booster BNT162b2 dose increases neutralizing antibody levels^3^. A phase 3 randomized controlled trial conducted in the delta (B.1.617.2) variant era reported fewer SARS-CoV-2 infections in persons randomized to a booster dose of BNT162b2 (6 out of 5081) than in persons randomized to placebo (123 out of 5044)^4^. Real-world studies conducted in the delta variant era estimated that booster vaccine effectiveness (VE) against infection, hospitalization and death was very high ranging from 86 to 95.3%^5–10^.

In December 2021, the omicron (B.1.1.529) variant became dominant worldwide, including in the United States^11^. The omicron variant is thought to be more likely to evade immunity or cause breakthrough infection after vaccination than the delta variant^12^. Indeed, emerging evidence suggests that booster VE against infection and hospitalization caused by the omicron variant is much lower^9, 13–15^. Booster VE against the omicron variant requires further investigation especially in older, racially/ethnically diverse populations with high prevalence of comorbid conditions, who have the highest risk of morbidity and mortality from COVID-19. Furthermore, studies are needed to determine whether booster VE varies by type of booster, type of primary vaccine, time since primary vaccine, age group and comorbidity burden in the omicron era.

The Veterans Affairs (VA) healthcare system, the largest national, integrated healthcare system in the United States, includes a large proportion of older adults with multimorbidity and has provided an adequate framework for multiple target trial emulation studies of the comparative effectiveness of COVID-19 vaccination^16–18^. We used target trial emulation principles^19^ to specify and emulate a trial comparing booster mRNA COVID-19 vaccination versus no booster during the omicron era with respect to risk of infection, hospitalization and death.

## Methods

### Specification and emulation of target trials: Overall study design

We designed this retrospective matched cohort study to emulate a target randomized controlled trial of booster (i.e. 3^rd^ dose) mRNA COVID-19 vaccination (BNT162b2 or mRNA-1273) versus no booster among persons who had already received two mRNA COVID-19 vaccinations at least 5 months earlier in the national VA healthcare system in a period of omicron variant predominance. The enrollment and follow-up period was between 12/1/2021 and 03/31/2022. During this period all persons who had completed more than 5 months since their primary COVID-19 immunization were eligible for booster vaccination in the VA irrespective of age or risk factors. We used a matched cohort design to emulate the balance achieved through randomization, with eligible persons matched on the date of their booster dose to their comparator(s) who had not received a booster dose as of that date. Each matched set was followed from the date of booster vaccination (“time zero”). This design ensured that the same time zero served as the date at which eligibility was determined and treatment was initiated and anchored the follow-up for both study arms (**Supplementary Figure 1**), thus minimizing immortal time and selection biases^20^. For each participant follow-up continued until the earliest of the following events: occurrence of an outcome event, death unrelated to COVID-19, booster vaccination of the comparator (with matched set censoring), or the end of the follow-up period (March 31, 2022). **Supplementary Table 1** compares the critical study design features of the specified and emulated target trials.

We used data from the VA’s Corporate Data Warehouse (CDW), a relational database of VA enrollees’ comprehensive electronic health records (EHR) and the VA COVID-19 Shared Data Resource (CSDR), which includes analytic variables provisioned by the VA Informatics and Computing Infrastructure (VINCI)^21^. We supplemented with additional claims data from the VA Community Care program (non-VA care paid by a VA facility) and from the Centers for Medicare and Medicaid Services (CMS) obtained through the VA Information Resource Center (VIReC)^22^.

### Eligibility criteria and study population

We identified all VA enrollees aged 18 years or older who were alive as of 12/01/2021 and had received exactly two doses of mRNA vaccination documented in the VA system with the second dose administered at least 5 months earlier (n=2,097,357) – see **Figure 1**. We excluded persons who: had evidence of SARS-CoV-2 infection prior to 12/01/2021, using a combination of VA and CMS data; had incongruent first and second doses of primary vaccination; had missing residential address zip code; did not have an inpatient or outpatient or primary care encounter in the VA healthcare system in the preceding 24 months; or were not assigned to a unique VA Integrated Service Network (VISN), the 19 administrative regions of VA^23^. Among the remaining 1,687,421 persons, 505,585 received a booster mRNA vaccine dose between 12/01/2021 and 03/31/2022 documented in the VA healthcare system and could potentially be included in the booster arm of the emulated trial if they could be matched to appropriate unboosted comparators. We captured booster doses administered by VA pharmacy, through VA-funded community care, outside the VA and documented in VA records or documented in CMS data.

**Figure 1.**
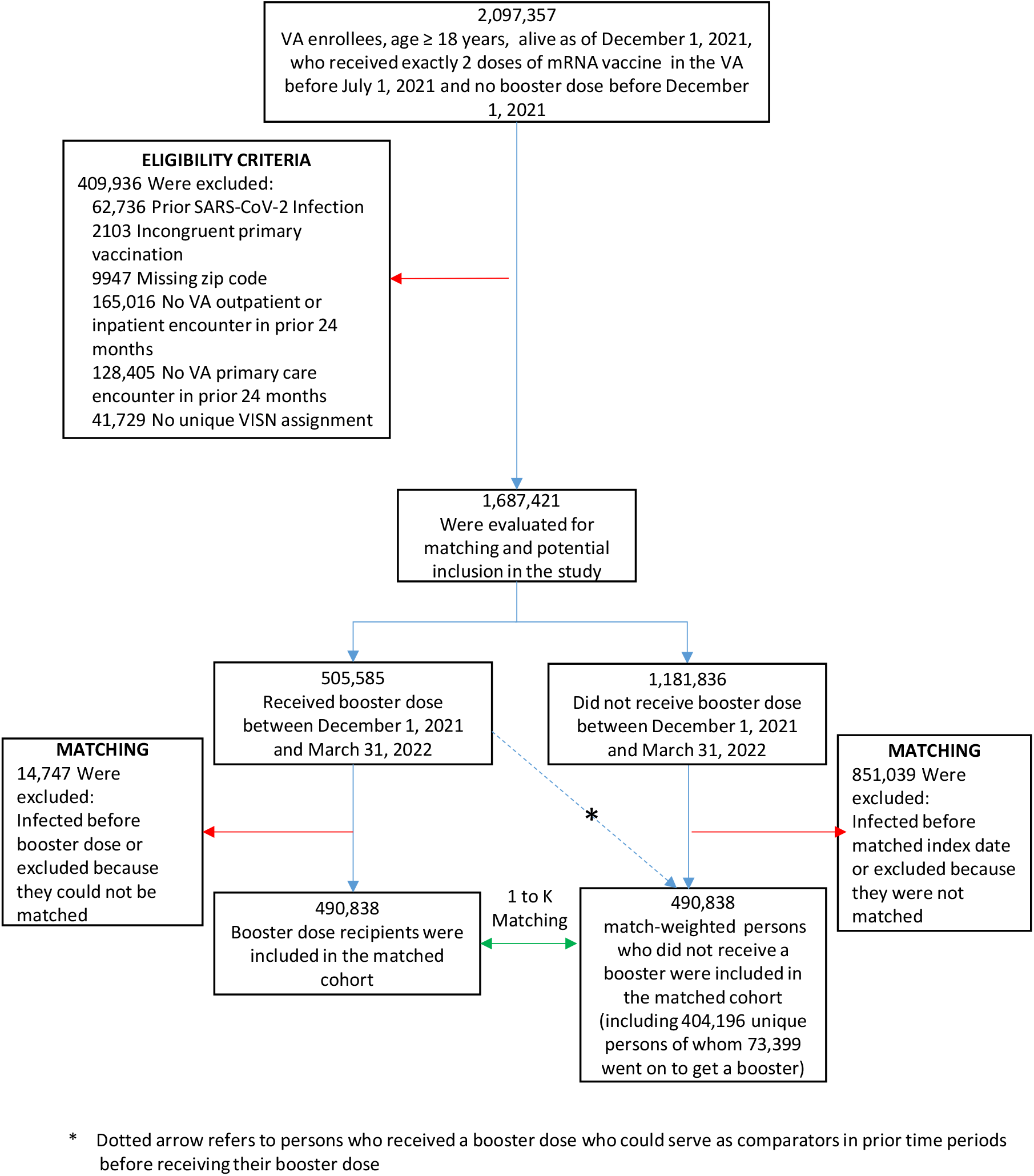
Eligibility criteria and matching process resulting in the selection of study populations for the emulation of a target trial comparing the effectiveness of booster mRNA COVID-19 vaccination versus no booster in the national Veterans Affairs healthcare system from December 1, 2021 to March 31, 2022.

### Cohort matching

We matched eligible persons who did or did not receive a booster anchored around the same calendar date of booster vaccination^17, 24^. In each 1-week period of the study, we identified among the eligible study participants those who were alive, uninfected and had not yet received booster vaccination as of the first day of the week. Among them, we identified all potential participants in the treatment arm, defined as those who received a booster mRNA vaccine during the given week and all potential comparators, who had not received a booster vaccine at the start of the week. We used the baseline characteristics of all persons to identify the best-matched comparator(s) for each person who received a booster dose using exact matching based on VISN, age (6-year buckets), Charlson Comorbidity Index^25^ (CCI) (3-point buckets), primary COVID-19 vaccination type (BNT162b2 or mRNA-1273) and date of completion of primary vaccination (6-week buckets). The exact matching factors were selected because they were known to be strongly associated with the probability of receiving a booster (the exposure) as well as with the risk of development of SARS-CoV-2 infection, hospitalization or death (the outcomes) in VA patients^26–30^.

To further reduce confounding as numerous covariates not used in the match remained imbalanced after exact matching, we then executed propensity score matching (calipers within 0.2 standard deviations from the mean) with replacement to identify the best match for each booster dose recipient in a 1:K variable ratio, where K varied based on the number of propensity score ties. We used the following covariates in the propensity score logistic regression model selected *a priori* based on being associated with both the exposure and the outcome: age, sex, self-reported race and ethnicity, urban/rural residence (based on zip codes, using data from the VA Office of Rural Health^31^, which uses the Secondary Rural-Urban Commuting Area [RUCA] for defining rurality), CCI, body mass index (BMI, calculated using measured weight and height), diabetes, congestive heart failure (CHF), chronic obstructive pulmonary disease (COPD), chronic kidney disease (CKD), receipt of immunosuppressant medications in the 2 years prior (see **Supplementary Table 2**) and the Care Assessment Need (CAN) score. The CAN score is a validated measure of 1-year mortality in VA enrollees calculated using socio-demographics, clinical diagnoses, vital signs, medications, laboratory values, and health care utilization data from VA’s national EHR^26, 32^. Diabetes, CHF, COPD and CKD were defined by international classification of disease, tenth revision (ICD-10) codes documented in VA EHR in the 2-year period prior to the study(see **Supplementary Table 3** for these ICD10 codes)^33^. The two-step matching process was repeated 16 times for each 1-week period between 12/01/21 and 03/31/22, with a separate propensity score model developed for each week to account for dynamic changes in booster uptake and infection incidence that were occurring during the study period. Persons who received booster vaccination were eligible for inclusion in the treatment arm, even if they had previously been selected as a matched comparator. Persons who did not receive a booster dose could serve as matched comparators to more than one person in the treatment arm. The matching strategy was implemented using STATA’s kmatch command^34^ (StataCorp, College Station, TX. USA).

### Primary endpoints

The trial had three primary endpoints: SARS-CoV-2 infection, SARS-CoV-2-related hospitalization and SARS CoV-2-related death.

SARS-CoV-2 infection was defined as a positive test for SARS-CoV-2 nucleic acid amplification test or antigen in a respiratory specimen within the VA system as well as such tests performed outside the VA but documented in VA records. Such positive tests are identified by the VA National Surveillance Tool and have been used by our group and others to support target trial emulation studies^16–18^. The earliest date of a documented positive test was taken as each participant’s date of infection.

SARS-CoV-2-related hospitalization was defined as hospitalization documented in the VA healthcare system or in CMS on or within 30 days after a positive test and SARS-CoV-2-related death as death from any cause within 30 days of a positive test; these definitions have been used in multiple prior VA studies^16–18, 26, 27, 29, 30^. Deaths occurring both within and outside the VA are comprehensively captured in the VA Corporate Data Warehouse (CDW) from a variety of VA and non-VA sources including VA inpatient files, VA Beneficiary Identification and Records Locator System (BIRLS), Social Security Administration (SSA) death files, and the Department of Defense ^35^. To ensure that deaths and hospitalizations that occurred on or before 03/31/22 were adequately captured we last accessed relevant data sources on 05/21/22.

### Statistical analysis: estimates of booster vaccine effectiveness

Baseline characteristics between the booster and no booster groups were compared using standardized differences. Unadjusted differences in incident outcome rates were estimated via Kaplan-Meier estimator for a period of 121 days, the study’s maximum follow-up.

We used Cox proportional hazards regression to compare booster vaccine recipients and their matched comparators with respect to the risk of developing the three study outcomes, with follow-up extending to 03/31/2022. Follow-up began from time zero in both groups and was limited to matched sets in which all individuals were still at risk 10 days after time zero since there is no expectation of protective effect within the first 10 days after vaccination. We calculated the incidence of each outcome and the hazard ratio adjusted for type of primary COVID-19 mRNA vaccine, time since 2^nd^ COVID-19 mRNA vaccine, sex, age, race, ethnicity, urban/rural residence, BMI, CCI, diabetes, CKD, CHF, COPD, CAN score, and immunosuppressant medications and stratified by VA VISN, to account for any residual confounding that might have been present after matching. Booster vaccine effectiveness (VE) was estimated as (1-adjusted hazard ratio) and served as the study’s primary outcome. Booster VE was estimated for the entire population and also for subgroups determined *a priori* based on type of booster (BNT162b2 or mRNA-1273), type of primary vaccination (BNT162b2 or mRNA-1273), time since primary vaccination (5-9 or >9 months), age groups, CCI categories. A χ^2^-test of no difference between the predicted adjusted hazard ratios in models with and without an interaction term was used to evaluate differences in VE between subgroups.

Missing values in sex, BMI, CAN score, race and ethnicity were uncommon and were imputed using deterministic imputation.

We compared as a negative outcome control^36^ the incidence of SARS-CoV-2 infection within ten days after time zero in the two groups, which should not be affected by vaccination, to verify there was no uncontrolled residual confounding after matching or any ascertainment bias.

All analyses were weighted to account for variable-ratio matching and matching with replacement. A robust sandwich-type variance estimator was used to account for clustering within matched group (because of ties in the propensity score), clustering within subjects (because of matching with replacement), and clustering in the cross-classification of the matched and within subject clusters^37^. We verified that the proportional hazards assumption was met using log-log plots and Schoenfeld residuals.

## Results

### Baseline characteristics of emulated trial participants

Baseline characteristics were well balanced between the booster arm (n=490,838) and their matched comparators (n=490,838 representing 404,196 unique persons) (**Table 1**). Matching with replacement allowed the matching of >97% of persons who received a booster dose. Baseline characteristics of all eligible boosted and unboosted individuals before matching are shown in **Supplementary Table 4** and **Supplementary Figure 2**.

**Table 1.**
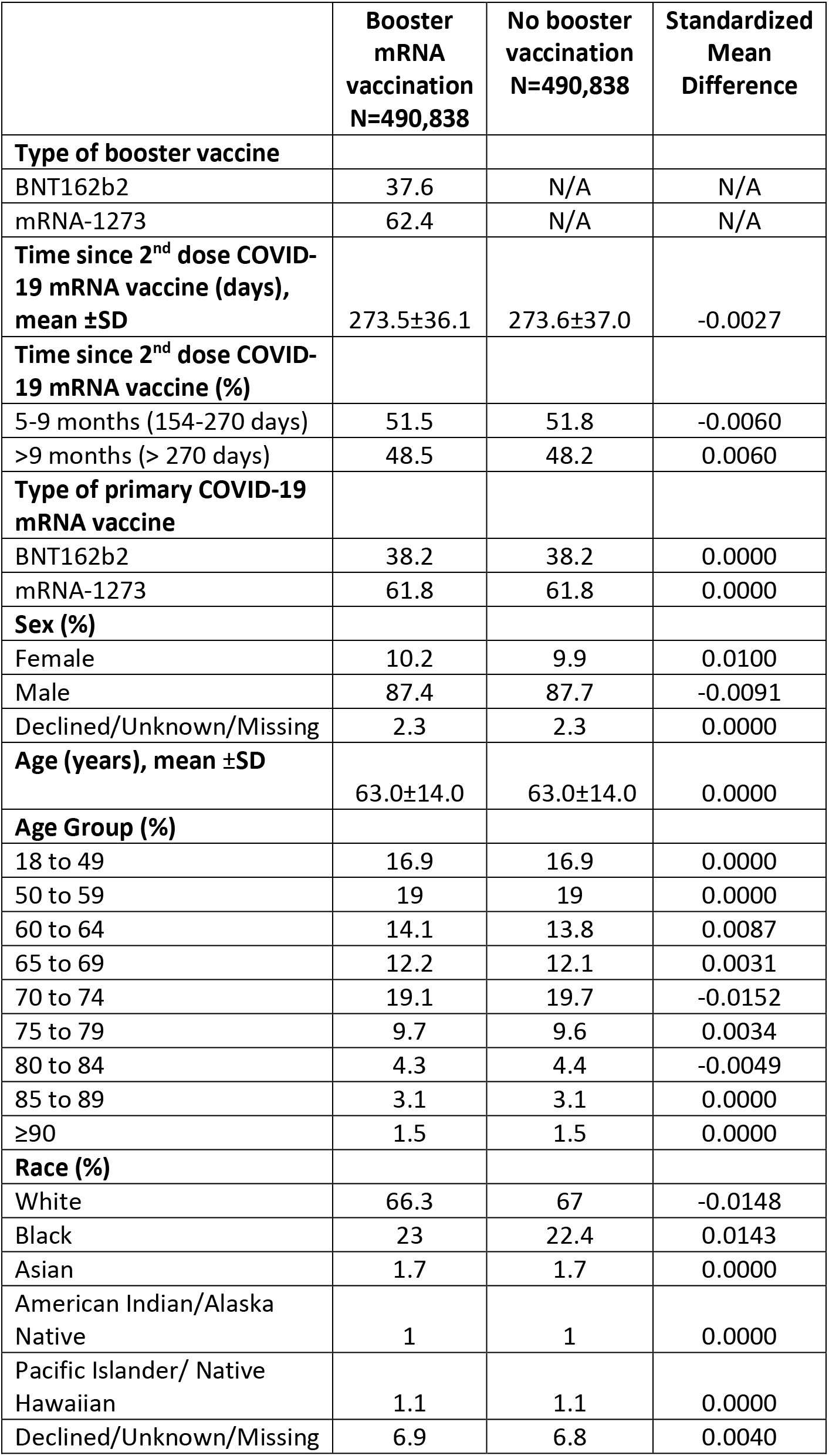
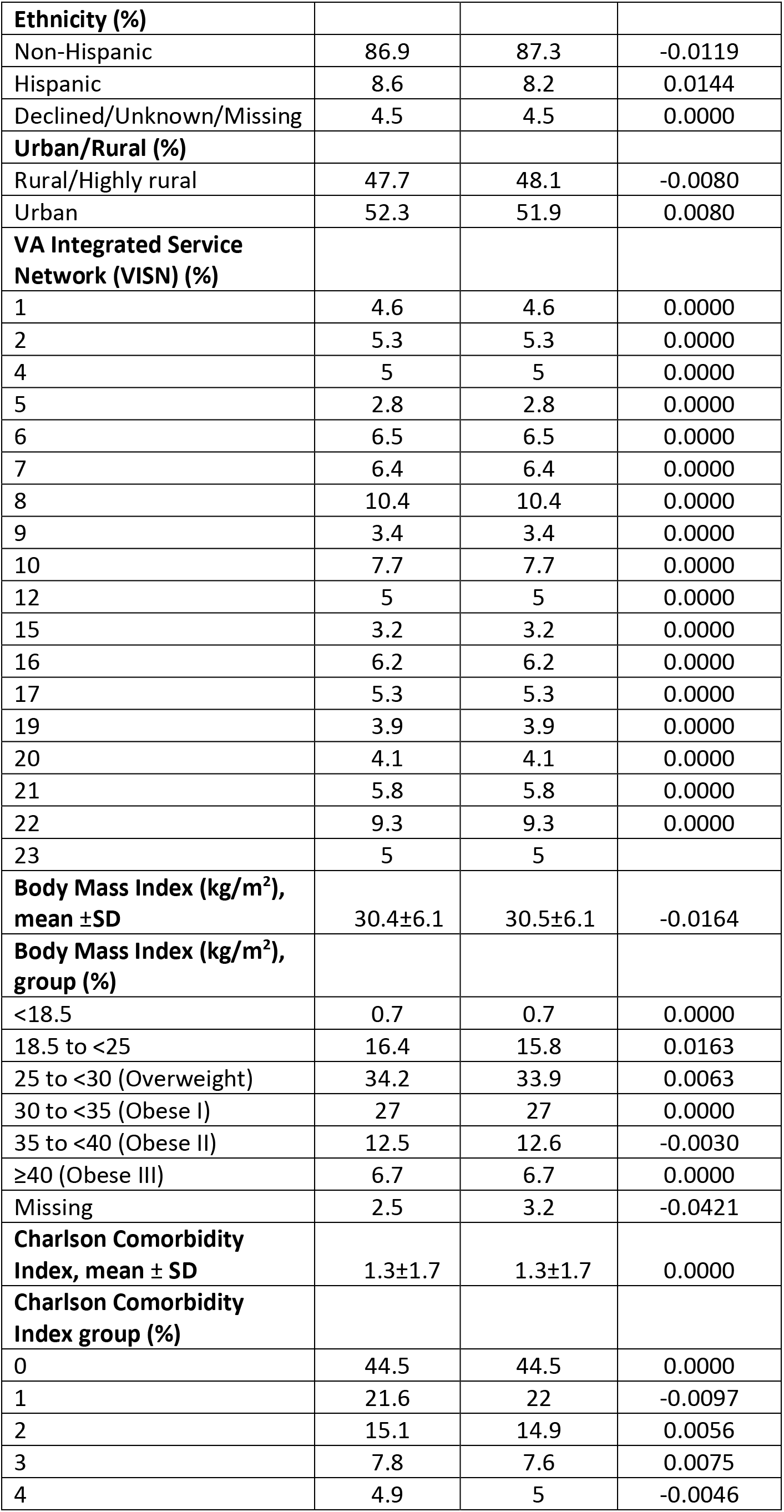
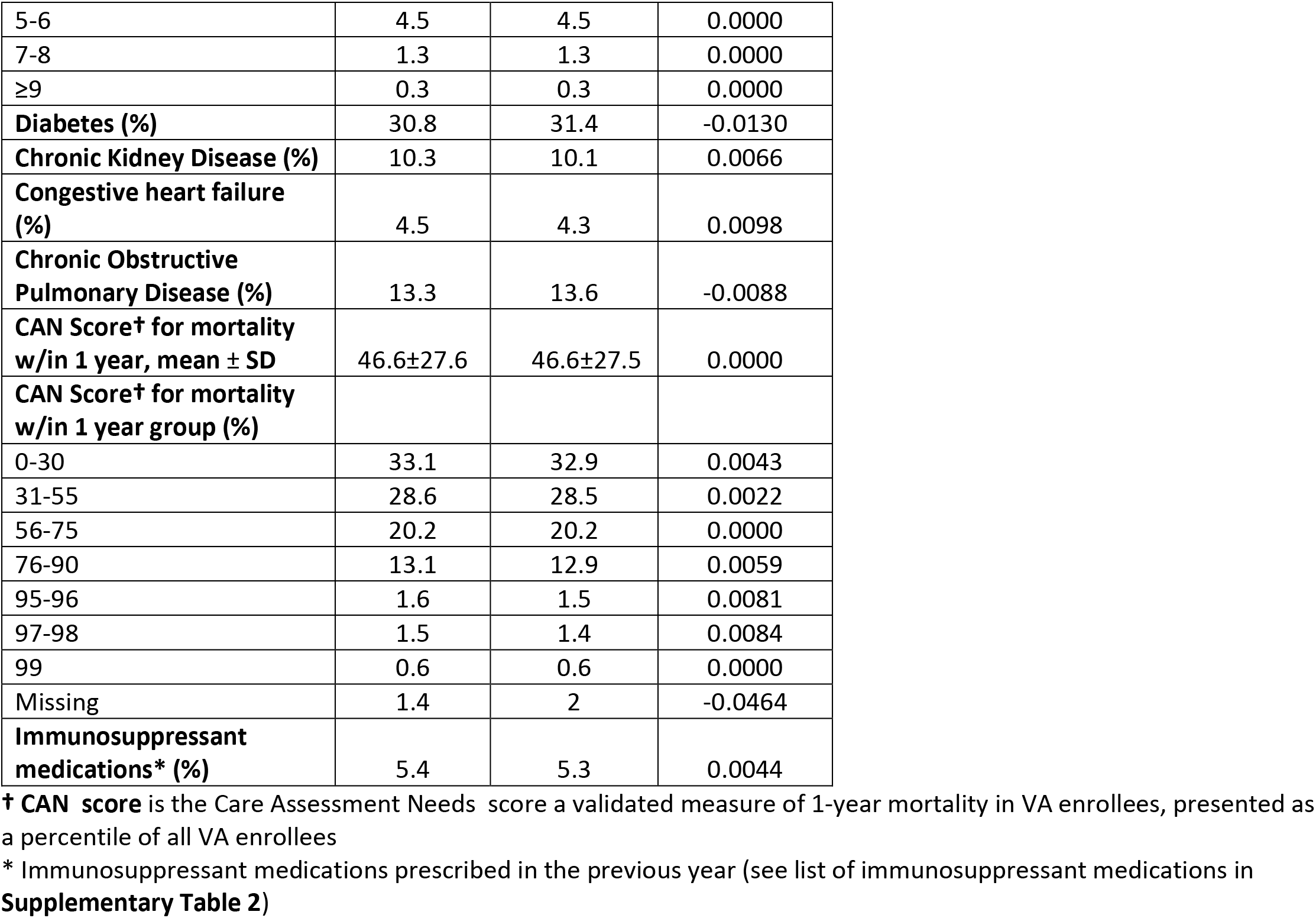
Baseline sociodemographic and clinical characteristics of persons who received a booster dose of mRNA COVID-19 vaccination in the VA healthcare system and their matched counterparts who did not receive a booster dose from December 1, 2021 to March 31, 2022.

|  | **Booster mRNA vaccination**<br>**N=490,838** | **No booster vaccination**<br>**N=490,838** | **Standardized Mean Difference** |
| --- | --- | --- | --- |
| **Type of booster vaccine** |  |  |  |
| BNT162b2 | 37.6 | N/A | N/A |
| mRNA-1273 | 62.4 | N/A | N/A |
| **Time since 2<sup>nd</sup> dose COVID-19 mRNA vaccine (days), mean ±SD** | 273.5±36.1 | 273.6±37.0 | -0.0027 |
| **Time since 2<sup>nd</sup> dose COVID-19 mRNA vaccine (%)** |  |  |  |
| 5-9 months (154-270 days) | 51.5 | 51.8 | -0.0060 |
| >9 months (> 270 days) | 48.5 | 48.2 | 0.0060 |
| **Type of primary COVID-19 mRNA vaccine** |  |  |  |
| BNT162b2 | 38.2 | 38.2 | 0.0000 |
| mRNA-1273 | 61.8 | 61.8 | 0.0000 |
| **Sex (%)** |  |  |  |
| Female | 10.2 | 9.9 | 0.0100 |
| Male | 87.4 | 87.7 | -0.0091 |
| Declined/Unknown/Missing | 2.3 | 2.3 | 0.0000 |
| **Age (years), mean ±SD** | 63.0±14.0 | 63.0±14.0 | 0.0000 |
| **Age Group (%)** |  |  |  |
| 18 to 49 | 16.9 | 16.9 | 0.0000 |
| 50 to 59 | 19 | 19 | 0.0000 |
| 60 to 64 | 14.1 | 13.8 | 0.0087 |
| 65 to 69 | 12.2 | 12.1 | 0.0031 |
| 70 to 74 | 19.1 | 19.7 | -0.0152 |
| 75 to 79 | 9.7 | 9.6 | 0.0034 |
| 80 to 84 | 4.3 | 4.4 | -0.0049 |
| 85 to 89 | 3.1 | 3.1 | 0.0000 |
| ≥90 | 1.5 | 1.5 | 0.0000 |
| **Race (%)** |  |  |  |
| White | 66.3 | 67 | -0.0148 |
| Black | 23 | 22.4 | 0.0143 |
| Asian | 1.7 | 1.7 | 0.0000 |
| American Indian/Alaska Native | 1 | 1 | 0.0000 |
| Pacific Islander/ Native Hawaiian | 1.1 | 1.1 | 0.0000 |
| Declined/Unknown/Missing | 6.9 | 6.8 | 0.0040 |
| **Ethnicity (%)** |  |  |  |
| Non-Hispanic | 86.9 | 87.3 | -0.0119 |
| Hispanic | 8.6 | 8.2 | 0.0144 |
| Declined/Unknown/Missing | 4.5 | 4.5 | 0.0000 |
| **Urban/Rural (%)** |  |  |  |
| Rural/Highly rural | 47.7 | 48.1 | -0.0080 |
| Urban | 52.3 | 51.9 | 0.0080 |
| **VA Integrated Service Network (VISN) (%)** |  |  |  |
| 1 | 4.6 | 4.6 | 0.0000 |
| 2 | 5.3 | 5.3 | 0.0000 |
| 4 | 5 | 5 | 0.0000 |
| 5 | 2.8 | 2.8 | 0.0000 |
| 6 | 6.5 | 6.5 | 0.0000 |
| 7 | 6.4 | 6.4 | 0.0000 |
| 8 | 10.4 | 10.4 | 0.0000 |
| 9 | 3.4 | 3.4 | 0.0000 |
| 10 | 7.7 | 7.7 | 0.0000 |
| 12 | 5 | 5 | 0.0000 |
| 15 | 3.2 | 3.2 | 0.0000 |
| 16 | 6.2 | 6.2 | 0.0000 |
| 17 | 5.3 | 5.3 | 0.0000 |
| 19 | 3.9 | 3.9 | 0.0000 |
| 20 | 4.1 | 4.1 | 0.0000 |
| 21 | 5.8 | 5.8 | 0.0000 |
| 22 | 9.3 | 9.3 | 0.0000 |
| 23 | 5 | 5 |  |
| **Body Mass Index (kg/m<sup>2</sup>), mean ±SD** | 30.4±6.1 | 30.5±6.1 | -0.0164 |
| **Body Mass Index (kg/m<sup>2</sup>), group (%)** |  |  |  |
| <18.5 | 0.7 | 0.7 | 0.0000 |
| 18.5 to <25 | 16.4 | 15.8 | 0.0163 |
| 25 to <30 (Overweight) | 34.2 | 33.9 | 0.0063 |
| 30 to <35 (Obese I) | 27 | 27 | 0.0000 |
| 35 to <40 (Obese II) | 12.5 | 12.6 | -0.0030 |
| ≥40 (Obese III) | 6.7 | 6.7 | 0.0000 |
| Missing | 2.5 | 3.2 | -0.0421 |
| **Charlson Comorbidity Index, mean ± SD** | 1.3±1.7 | 1.3±1.7 | 0.0000 |
| **Charlson Comorbidity Index group (%)** |  |  |  |
| 0 | 44.5 | 44.5 | 0.0000 |
| 1 | 21.6 | 22 | -0.0097 |
| 2 | 15.1 | 14.9 | 0.0056 |
| 3 | 7.8 | 7.6 | 0.0075 |
| 4 | 4.9 | 5 | -0.0046 |
| 5-6 | 4.5 | 4.5 | 0.0000 |
| 7-8 | 1.3 | 1.3 | 0.0000 |
| ≥9 | 0.3 | 0.3 | 0.0000 |
| **Diabetes (%)** | 30.8 | 31.4 | -0.0130 |
| **Chronic Kidney Disease (%)** | 10.3 | 10.1 | 0.0066 |
| **Congestive heart failure (%)** | 4.5 | 4.3 | 0.0098 |
| **Chronic Obstructive Pulmonary Disease (%)** | 13.3 | 13.6 | -0.0088 |
| **CAN Score† for mortality w/in 1 year, mean ± SD** | 46.6±27.6 | 46.6±27.5 | 0.0000 |
| **CAN Score† for mortality w/in 1 year group (%)** |  |  |  |
| 0-30 | 33.1 | 32.9 | 0.0043 |
| 31-55 | 28.6 | 28.5 | 0.0022 |
| 56-75 | 20.2 | 20.2 | 0.0000 |
| 76-90 | 13.1 | 12.9 | 0.0059 |
| 95-96 | 1.6 | 1.5 | 0.0081 |
| 97-98 | 1.5 | 1.4 | 0.0084 |
| 99 | 0.6 | 0.6 | 0.0000 |
| Missing | 1.4 | 2 | -0.0464 |
| **Immunosuppressant medications\* (%)** | 5.4 | 5.3 | 0.0044 |

† **CAN score** is the Care Assessment Needs score a validated measure of 1-year mortality in VA enrollees, presented as a percentile of all VA enrollees
\* Immunosuppressant medications prescribed in the previous year (see list of immunosuppressant medications in **Supplementary Table 2**)

Both matched groups were predominantly male (87.4% vs. 87.7%), had advanced mean age (63.0 ±14 yrs in both groups), diverse racial/ethnic distribution (e.g. Black 21.8% vs. 22.2%, Hispanic 8.6% vs. 8.2%) and a substantial comorbidity burden (mean CCI 1.3±1.7 in both groups). Major comorbid conditions such as diabetes, CHF, COPD and CKD and exposure to immunosuppressant medications were common and nearly equally distributed in the two groups. The mean number of days (273±36) since completion of primary vaccination was almost identical in the two groups with ∼48% in each group having received the second dose of the vaccine >9 months prior to time zero.

### Booster vaccine effectiveness against infection

486,616 out of 490,838 (99.1%) matched sets remained under follow-up and at risk at day 10 after time zero and were included in VE estimations (**Tables 2-4**). During a mean follow-up of 79.8 days identical in the two groups, (maximum 121 days), 17,384 SARS-CoV-2 infections were documented >10 days after time zero, including 6,438 in the booster cohort and 10,946 in the matched no booster cohort (**Table 2**). 85% (14,823/17,384) of these infections occurred after 01/01/22, when omicron accounted for almost 100% of infections^11^ in the USA and an additional 14% (2467/17,384) from 12/16-12/31/22 when omicron accounted for the majority of infections, while only 0.5% (94/17,384) occurred before 12/16/22.

**Table 2.**
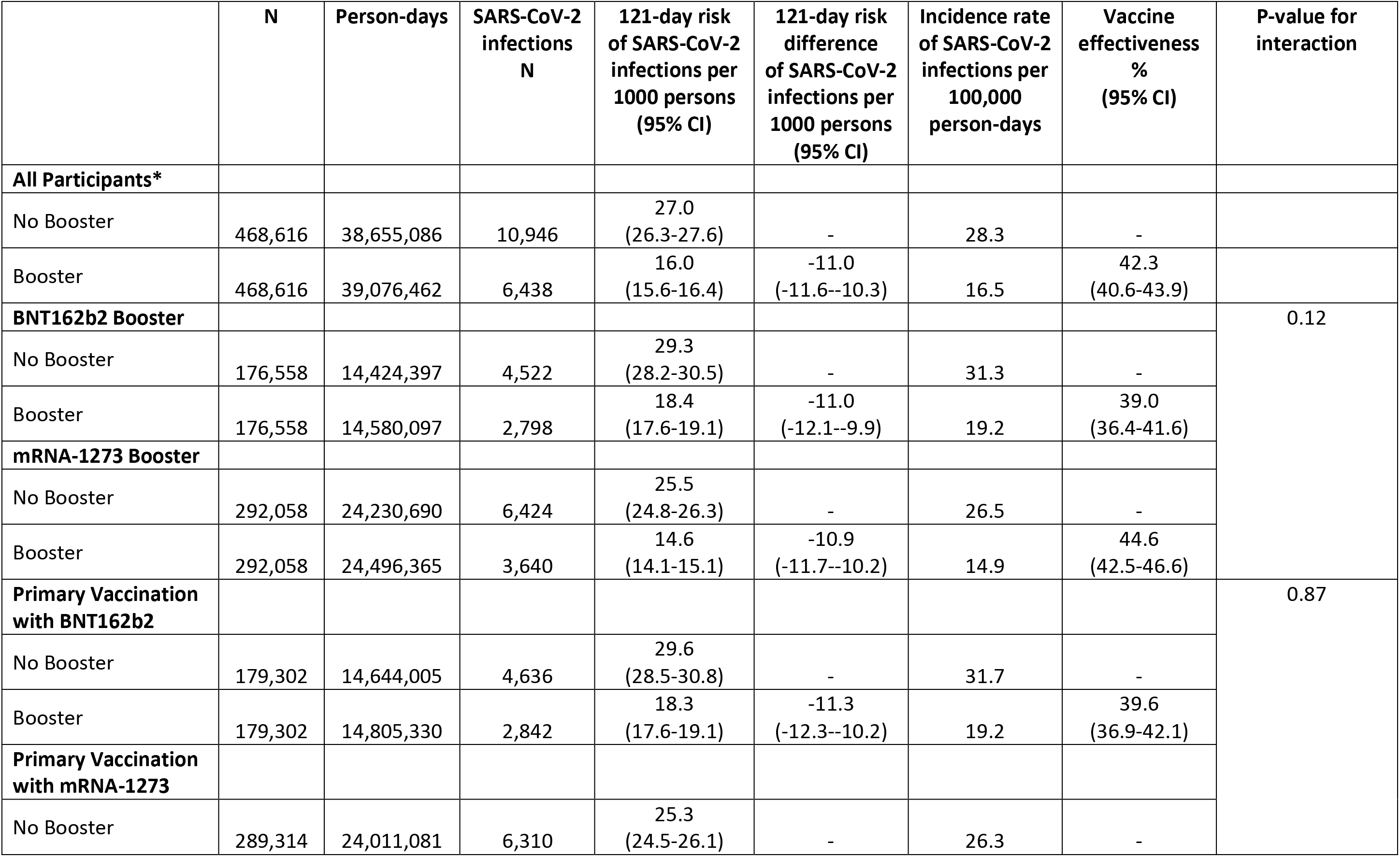
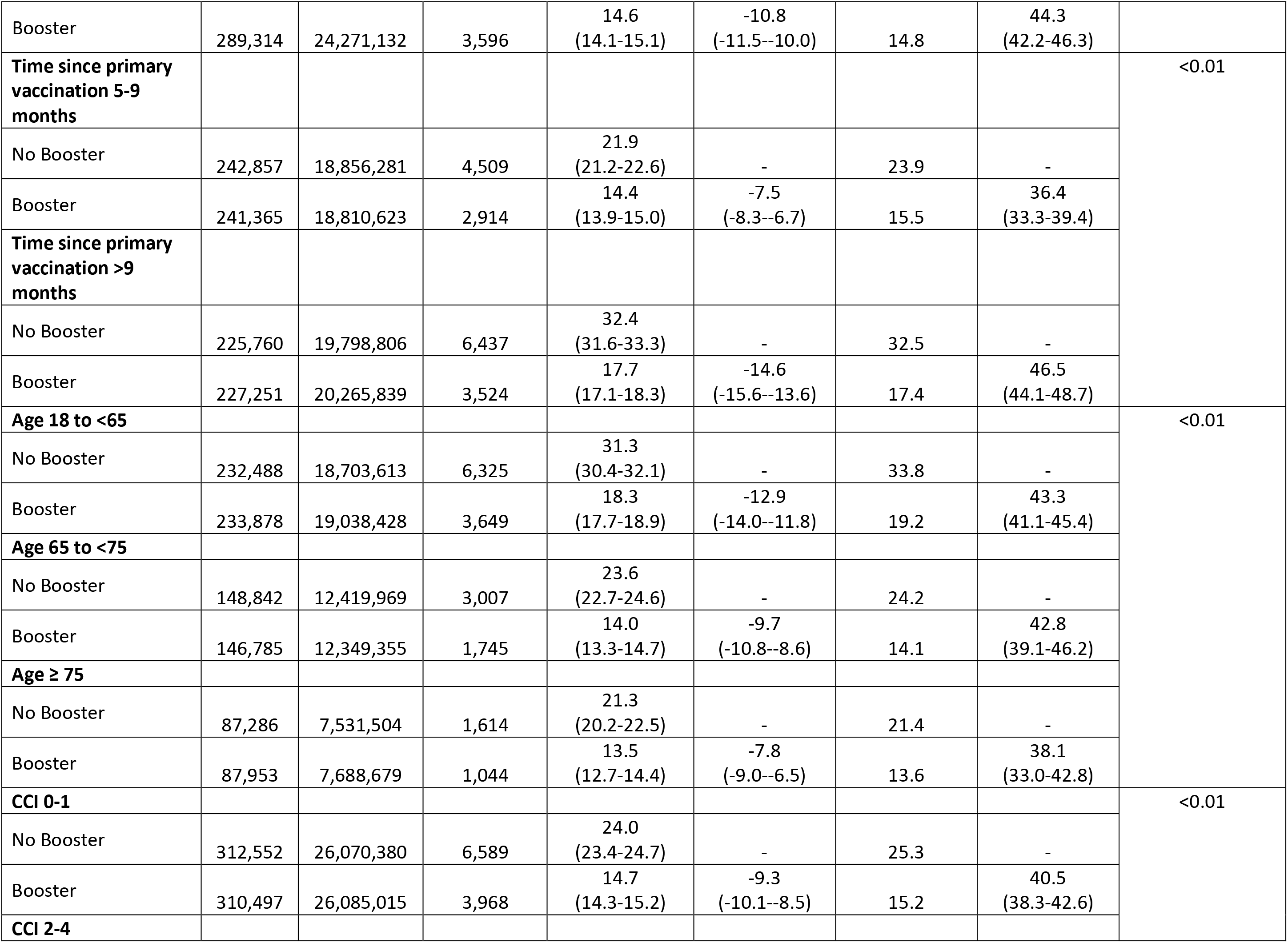
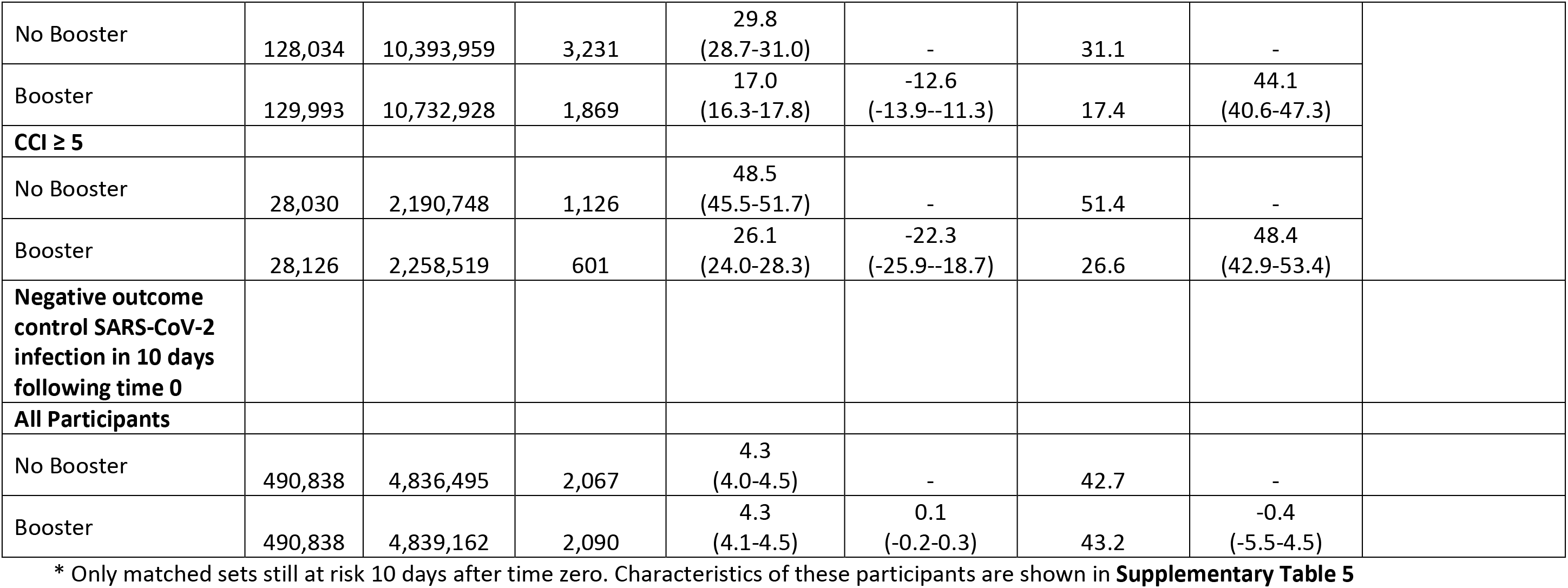
Comparison of persons who received a booster dose of mRNA COVID-19 vaccination in the VA healthcare system and their matched counterparts who did not receive a booster dose from December 1, 2021 to March 31, 2022 with respect to the risk of developing SARS-CoV-2 <u>infection</u> and estimation of vaccine effectiveness.

Cumulative incidence of infection at 121 days was significantly lower in the booster (16.0 per 1000 persons) than in the no booster (27.0 per 1000) cohort (risk difference -11 per 1000, 95% CI -11.6 to -10.3) (**Figure 2** **and Table 2**). Booster VE against infection was 42.3% (95% CI 40.6-43.9) overall, 39.0% (36.4-41.6) for BNT162b2 and 44.6% (42.5-46.6) for mRNA-1273 booster (interaction p-value 0.12) – see **Supplementary Figure 3** for cumulative incidence curves by booster type. Booster VE was higher in those with >9 months since primary vaccination (46.5% [44.1-48.5]) than in persons with 5-9 months since primary vaccination (36.4% [33.3-39.4]) (interaction p-value<0.01). Booster VE appeared to be higher in persons with more comorbidities (interaction p-value<0.01) but lower in older age groups(interaction p-value<0.01) (**Table 2**).

**Figure 2.**
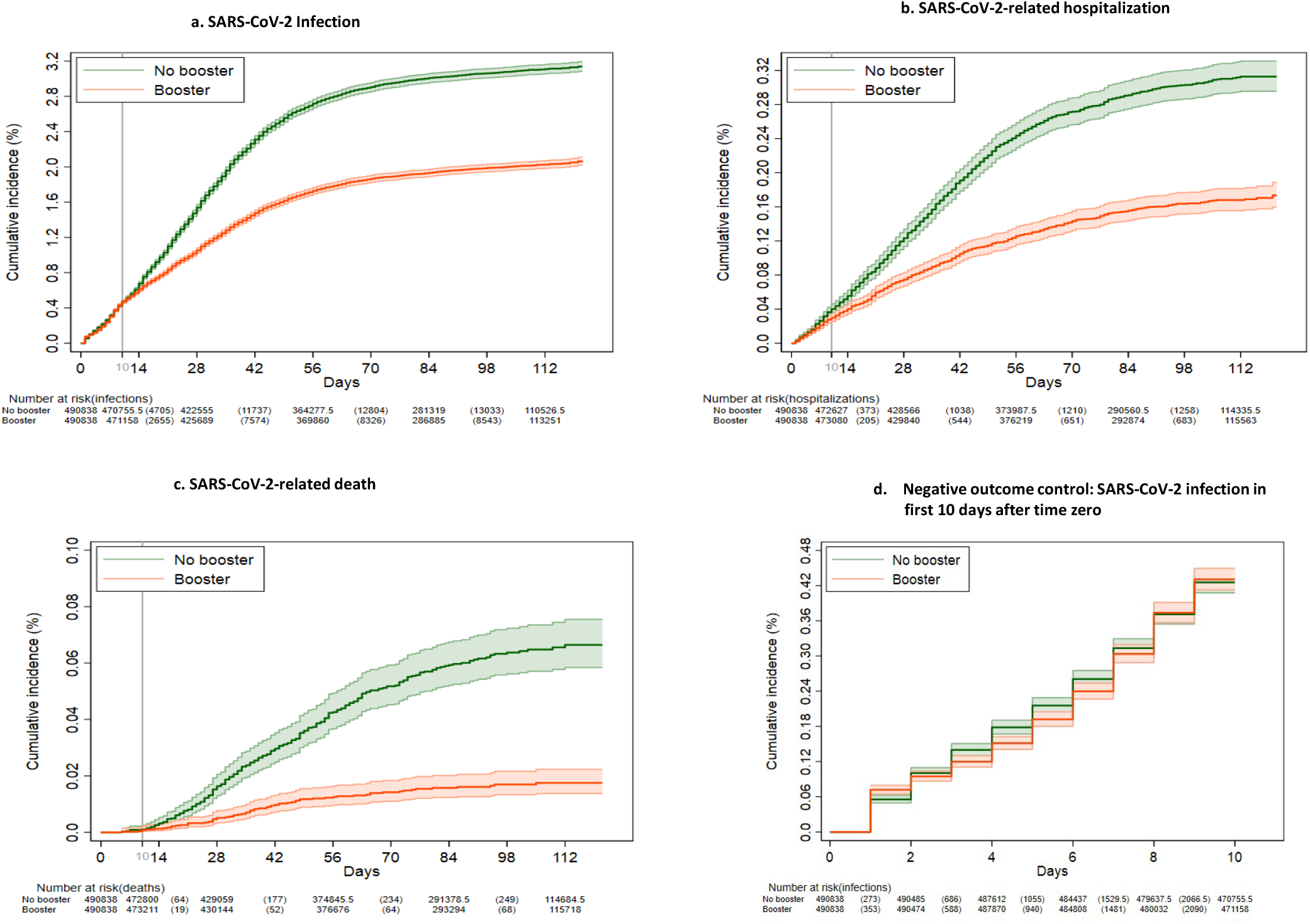
Kaplan-Meier curves comparing persons who received a booster mRNA COVID-19 vaccine versus their matched counterparts who did not with respect to the cumulative incidence (%) and 95% confidence intervals of SARS-CoV-2 infection (a), SARS-CoV-2-related hospitalization (b), SARS-CoV-2-related death (c) and SARS-CoV-2 infection during the first 10 days after time zero (d) (negative outcome control).

### Booster vaccine effectiveness against SARS-CoV-2-related hospitalization

A total of 1575 SARS-CoV-2-related hospitalizations were documented during follow-up, including 515 in the booster cohort and 1060 in the matched no booster cohort (**Table 3**). Cumulative incidence of SARS-CoV-2-related hospitalization at 121 days was significantly lower in the booster (1.3 per 1000 persons) than in the no booster (2.7 per 1000) group (risk difference -1.4 per 1000, 95% CI -1.6 to -1.2) (**Figure 2** **and Table 3**).

**Table 3.** Comparison of persons who received a booster dose of mRNA COVID-19 vaccination in the VA healthcare system and their matched counterparts who did not receive a booster dose from December 1, 2021 to March 31, 2022 with respect to the risk of developing SARS-CoV-2-related hospitalization and estimation of vaccine effectiveness.

|  | N | Person-days | SARS-CoV-2 Hospitalizations N | 121-day risk of SARS-CoV-2 hospitalizations per 1000 persons (95% CI) | 121-day risk difference of SARS-CoV-2 hospitalizations per 1000 persons (95% CI) | Incidence rate of SARS-CoV-2 hospitalizations per 100,000 person-days | Vaccine effectiveness % (95% CI) | P-value for interaction |
| --- | --- | --- | --- | --- | --- | --- | --- | --- |
| **All Participants\*** |  |  |  |  |  |  |  |  |
| No Booster | 468,616 | 39,305,524 | 1,060 | 2.7<br>(2.6-2.9) | - | 2.7 | - |  |
| Booster | 468,616 | 39,457,502 | 515 | 1.3<br>(1.2-1.5) | -1.4<br>(-1.6--1.2) | 1.3 | 53.3<br>(48.1-58.0) |  |
| **BNT162b2 Booster** |  |  |  |  |  |  |  | 0.59 |
| No Booster | 176,558 | 14,695,971 | 485 | 3.3<br>(3.0-3.7) | - | 3.3 | - |  |
| Booster | 176,558 | 14,748,878 | 232 | 1.6<br>(1.4-1.8) | -1.7<br>(-2.1--1.4) | 1.6 | 54.0<br>(46.1-60.8) |  |
| **mRNA-1273 Booster** |  |  |  |  |  |  |  |  |
| No Booster | 292,058 | 24,609,553 | 575 | 2.4<br>(2.2-2.6) | - | 2.3 | - |  |
| Booster | 292,058 | 24,708,624 | 283 | 1.2<br>(1.1-1.3) | -1.2<br>(-1.5--1.0) | 1.2 | 52.9<br>(45.6-59.2) |  |
| **Primary Vaccination with BNT162b2** |  |  |  |  |  |  |  | 0.91 |
| No Booster | 179,302 | 14,921,741 | 501 | 3.4<br>(3.0-3.7) | - | 3.4 | - |  |
| Booster | 179,302 | 14,976,081 | 241 | 1.6<br>(1.4-1.8) | -1.8<br>(-2.1--1.4) | 1.6 | 53.7<br>(45.8-60.4) |  |
| **Primary Vaccination with mRNA-1273** |  |  |  |  |  |  |  |  |
| No Booster | 289,314 | 24,383,783 | 559 | 2.3<br>(2.1-2.6) | - | 2.3 | - |  |
| Booster | 289,314 | 24,481,421 | 274 | 1.2<br>(1.0-1.3) | -1.2<br>(-1.4--0.9) | 1.1 | 53.1<br>(45.7-59.5) |  |
| **Time since primary vaccination 5-9 months** |  |  |  |  |  |  |  | <0.01 |
| No Booster | 242,857 | 19,107,546 | 535 | 2.7<br>(2.5-3.0) | - | 2.8 | - |  |
| Booster | 241,365 | 18,974,082 | 317 | 1.7<br>(1.5-1.8) | -1.1<br>(-1.4--0.8) | 1.7 | 43.8<br>(35.2-51.3) |  |
| **Time since primary vaccination >9 months** |  |  |  |  |  |  |  |  |
| No Booster | 225,760 | 20,197,978 | 525 | 2.7<br>(2.5-3.0) | - | 2.6 | - |  |
| Booster | 227,251 | 20,483,420 | 198 | 1.0<br>(0.9-1.2) | -1.7<br>(-2.0--1.4) | 1.0 | 63.2<br>(56.4-69.0) |  |
| **Age 18 to <65** |  |  |  |  |  |  |  | 0.29 |
| No Booster | 232,488 | 19,108,938 | 311 | 1.6<br>(1.4-1.8) | - | 1.6 | - |  |
| Booster | 233,878 | 19,265,332 | 149 | 0.8<br>(0.7-0.9) | -0.8<br>(-1.1--0.6) | 0.8 | 53.3<br>(43.2-61.6) |  |
| **Age 65 to <75** |  |  |  |  |  |  |  |  |
| No Booster | 148,842 | 12,585,729 | 422 | 3.5<br>(3.1-3.8) | - | 3.4 | - |  |
| Booster | 146,785 | 12,447,755 | 198 | 1.6<br>(1.4-1.9) | -1.8<br>(-2.2--1.4) | 1.6 | 54.2<br>(45.4-61.6) |  |
| **Age ≥ 75** |  |  |  |  |  |  |  |  |
| No Booster | 87,286 | 7,610,857 | 327 | 4.4<br>(3.9-4.9) | - | 4.3 | - |  |
| Booster | 87,953 | 7,744,415 | 168 | 2.2<br>(1.9-2.6) | -2.2<br>(-2.8--1.6) | 2.2 | 52.3<br>(42.1-60.7) |  |
| **CCI 0-1** |  |  |  |  |  |  |  | <0.01 |
| No Booster | 312,552 | 26,492,891 | 322 | 1.3<br>(1.1-1.4) | - | 1.2 | - |  |
| Booster | 310,497 | 26,334,294 | 163 | 0.6<br>(0.5-0.7) | -0.6<br>(-0.8--0.5) | 0.6 | 49.9<br>(39.2-58.7) |  |
| **CCI 2-4** |  |  |  |  |  |  |  |  |
| No Booster | 128,034 | 10,570,379 | 436 | 4.2<br>(3.8-4.6) | - | 4.1 | - |  |
| Booster | 129,993 | 10,836,389 | 213 | 2.0<br>(1.7-2.3) | -2.1<br>(-2.6--1.7) | 2.0 | 54.2<br>(45.7-61.4) |  |
| **CCI ≥ 5** |  |  |  |  |  |  |  |  |
| No Booster | 28,030 | 2,242,254 | 302 | 13.2<br>(11.7-14.9) | - | 13.5 | - |  |
| Booster | 28,126 | 2,286,819 | 139 | 6.2<br>(5.2-7.3) | -7.0<br>(-8.8--5.2) | 6.1 | 56.3<br>(46.4-64.3) |  |

\* Only matched sets still at risk 10 days after time zero. Characteristics of these participants are shown in **Supplementary Table 5**

Booster VE against hospitalization was 53.3% (48.1-58.0) overall, 54.0% (46.1-60.8) for BNT162b2 and 52.9% (45.6-59.2) for mRNA-1273 booster (p-value for interaction by booster type 0.59) – see **Supplementary Figure 3** for cumulative incidence curves by booster type. Booster VE was higher among persons with >9 months since primary vaccination (interaction p-value<0.01) and in persons with more comorbidities (interaction p-value<0.01) with no appreciable differences across age groups or primary vaccination types.

### Booster vaccine effectiveness against SARS-CoV-2-related death

A total of 262 SARS-CoV-2-related deaths were documented during follow-up, including 47 in the booster cohort and 215 in the matched no booster cohort (**Table 4**). Cumulative incidence of SARS-CoV-2-related death at 121 days was significantly lower in the booster (0.1 per 1000 persons) than in the no booster (0.6 per 1000) group (risk difference -0.5 per 1000, 95% CI -0.6 to -0.4) (**Figure 2** **and Table 4**). Booster VE against death was 79.1% (71.2-84.9) overall, 85.5% (73.9-92.0) for BNT162b2 and 75.2% (62.9-83.4) for mRNA-1273 booster (p-value for interaction by booster type 0.48) –see **Supplementary Figure 3** for cumulative incidence curves by booster type. Booster VE was not significantly different across age and CCI groups and according to time since primary vaccination or primary vaccination type.

**Table 4.**
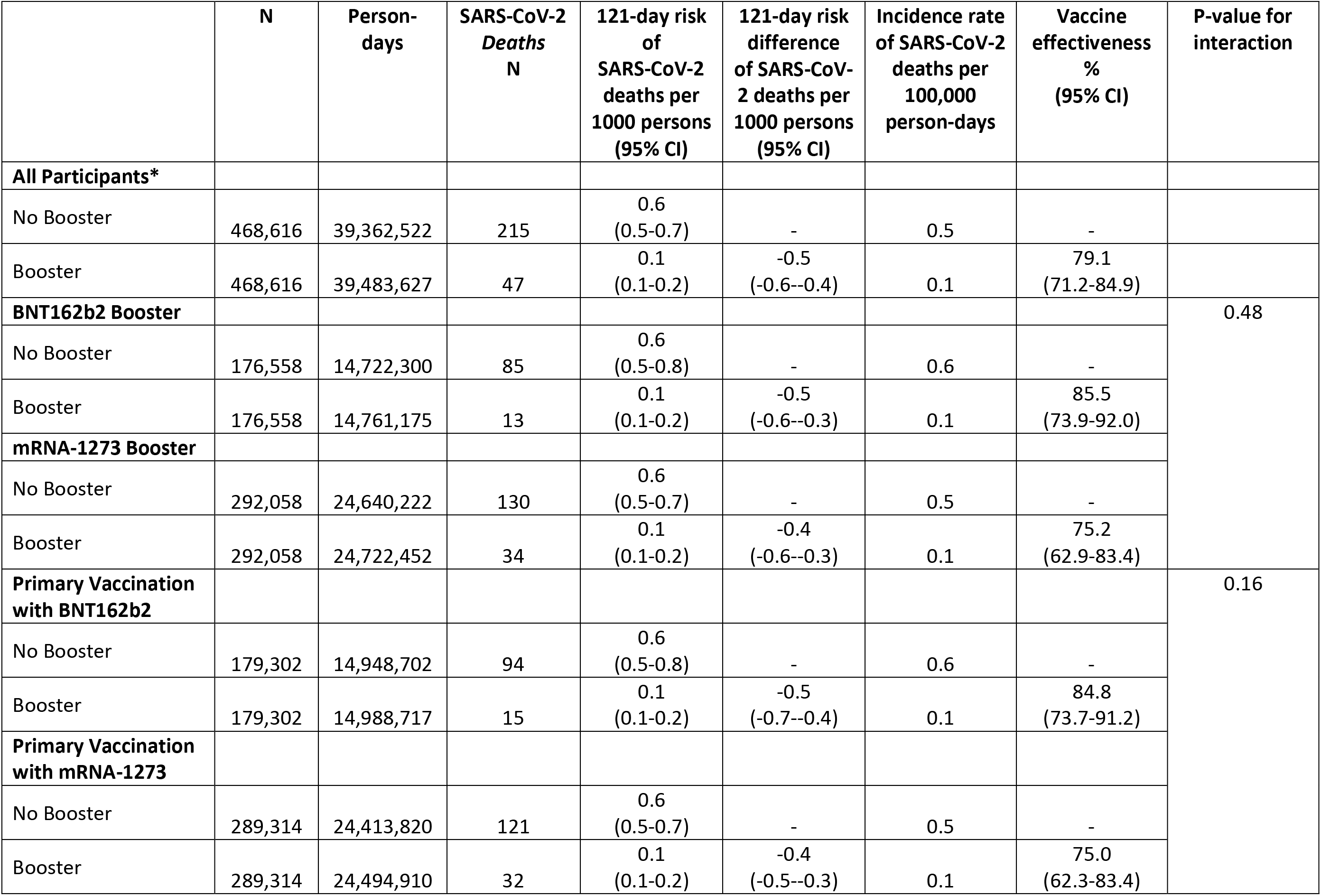
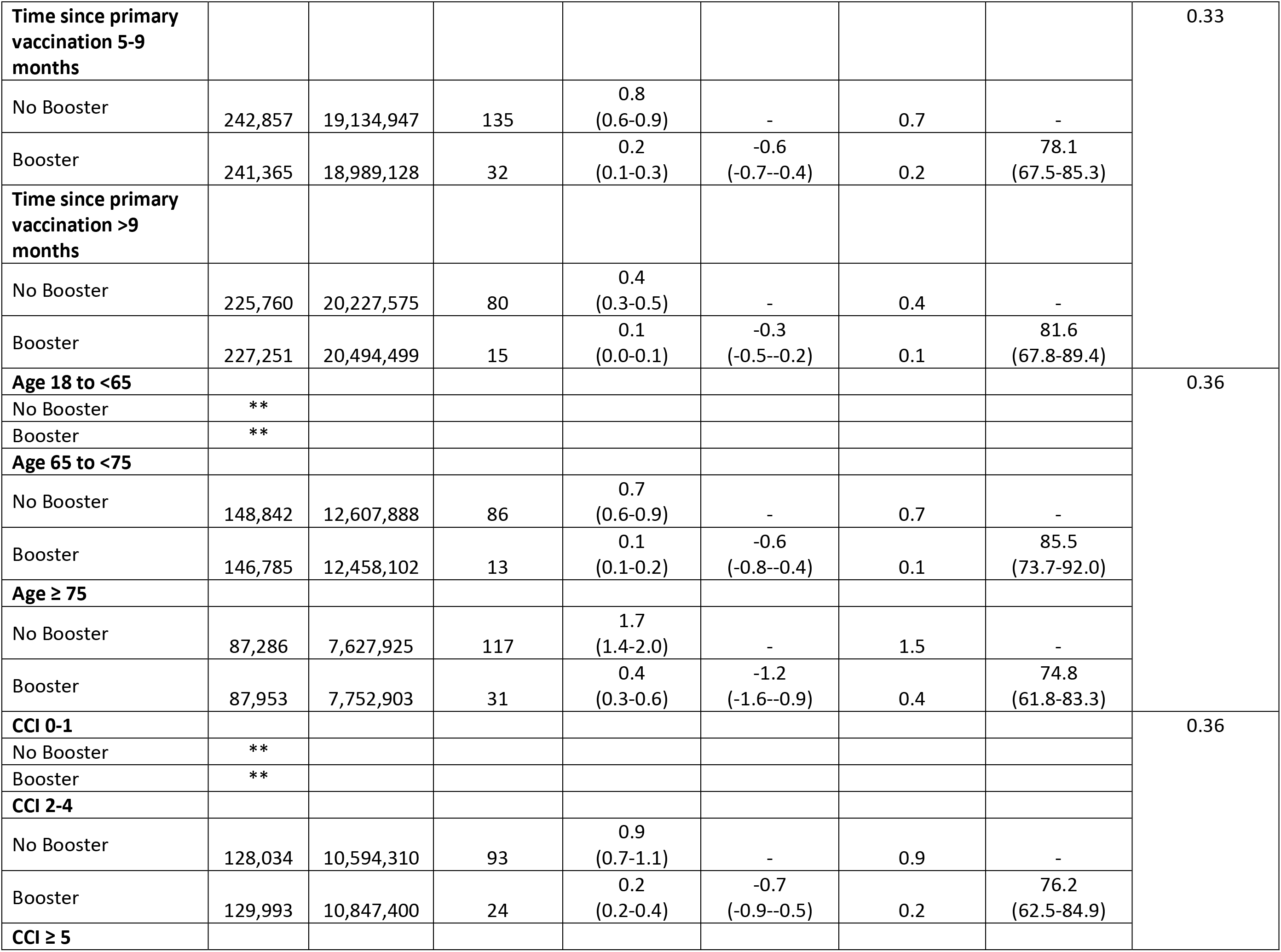
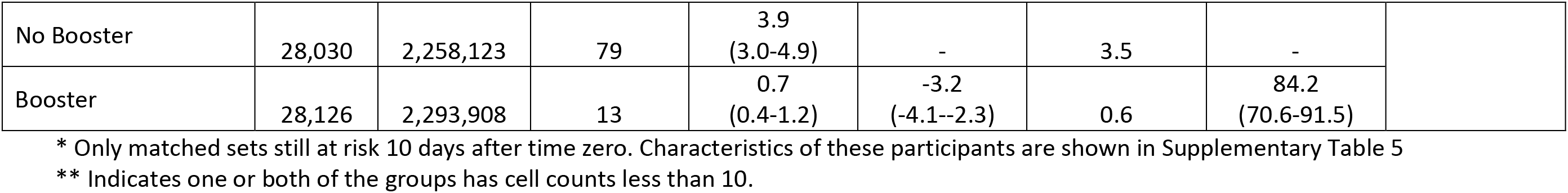
Comparison of persons who received a booster dose of mRNA COVID-19 vaccination in the VA healthcare system and their matched counterparts who did not receive a booster dose from December 1, 2021 to March 31, 2022 with respect to the risk of developing SARS-CoV-2-related death and estimation of vaccine effectiveness.

|  | N | Person-days | SARS-CoV-2 Deaths<br>N | 121-day risk of SARS-CoV-2 deaths per 1000 persons (95% CI) | 121-day risk difference of SARS-CoV-2 deaths per 1000 persons (95% CI) | Incidence rate of SARS-CoV-2 deaths per 100,000 person-days | Vaccine effectiveness % (95% CI) | P-value for interaction |
| --- | --- | --- | --- | --- | --- | --- | --- | --- |
| **All Participants\*** |  |  |  |  |  |  |  |  |
| No Booster | 468,616 | 39,362,522 | 215 | 0.6 (0.5-0.7) | - | 0.5 | - |  |
| Booster | 468,616 | 39,483,627 | 47 | 0.1 (0.1-0.2) | -0.5 (-0.6--0.4) | 0.1 | 79.1 (71.2-84.9) |  |
| **BNT162b2 Booster** |  |  |  |  |  |  |  | 0.48 |
| No Booster | 176,558 | 14,722,300 | 85 | 0.6 (0.5-0.8) | - | 0.6 | - |  |
| Booster | 176,558 | 14,761,175 | 13 | 0.1 (0.1-0.2) | -0.5 (-0.6--0.3) | 0.1 | 85.5 (73.9-92.0) |  |
| **mRNA-1273 Booster** |  |  |  |  |  |  |  |  |
| No Booster | 292,058 | 24,640,222 | 130 | 0.6 (0.5-0.7) | - | 0.5 | - |  |
| Booster | 292,058 | 24,722,452 | 34 | 0.1 (0.1-0.2) | -0.4 (-0.6--0.3) | 0.1 | 75.2 (62.9-83.4) |  |
| **Primary Vaccination with BNT162b2** |  |  |  |  |  |  |  | 0.16 |
| No Booster | 179,302 | 14,948,702 | 94 | 0.6 (0.5-0.8) | - | 0.6 | - |  |
| Booster | 179,302 | 14,988,717 | 15 | 0.1 (0.1-0.2) | -0.5 (-0.7--0.4) | 0.1 | 84.8 (73.7-91.2) |  |
| **Primary Vaccination with mRNA-1273** |  |  |  |  |  |  |  |  |
| No Booster | 289,314 | 24,413,820 | 121 | 0.6 (0.5-0.7) | - | 0.5 | - |  |
| Booster | 289,314 | 24,494,910 | 32 | 0.1 (0.1-0.2) | -0.4 (-0.5--0.3) | 0.1 | 75.0 (62.3-83.4) |  |
| **Time since primary vaccination 5-9 months** |  |  |  |  |  |  |  | 0.33 |
| No Booster | 242,857 | 19,134,947 | 135 | 0.8<br>(0.6-0.9) | - | 0.7 | - |  |
| Booster | 241,365 | 18,989,128 | 32 | 0.2<br>(0.1-0.3) | -0.6<br>(-0.7--0.4) | 0.2 | 78.1<br>(67.5-85.3) |  |
| **Time since primary vaccination >9 months** |  |  |  |  |  |  |  |  |
| No Booster | 225,760 | 20,227,575 | 80 | 0.4<br>(0.3-0.5) | - | 0.4 | - |  |
| Booster | 227,251 | 20,494,499 | 15 | 0.1<br>(0.0-0.1) | -0.3<br>(-0.5--0.2) | 0.1 | 81.6<br>(67.8-89.4) |  |
| **Age 18 to <65** |  |  |  |  |  |  |  | 0.36 |
| No Booster | ** |  |  |  |  |  |  |  |
| Booster | ** |  |  |  |  |  |  |  |
| **Age 65 to <75** |  |  |  |  |  |  |  |  |
| No Booster | 148,842 | 12,607,888 | 86 | 0.7<br>(0.6-0.9) | - | 0.7 | - |  |
| Booster | 146,785 | 12,458,102 | 13 | 0.1<br>(0.1-0.2) | -0.6<br>(-0.8--0.4) | 0.1 | 85.5<br>(73.7-92.0) |  |
| **Age ≥ 75** |  |  |  |  |  |  |  |  |
| No Booster | 87,286 | 7,627,925 | 117 | 1.7<br>(1.4-2.0) | - | 1.5 | - |  |
| Booster | 87,953 | 7,752,903 | 31 | 0.4<br>(0.3-0.6) | -1.2<br>(-1.6--0.9) | 0.4 | 74.8<br>(61.8-83.3) |  |
| **CCI 0-1** |  |  |  |  |  |  |  | 0.36 |
| No Booster | ** |  |  |  |  |  |  |  |
| Booster | ** |  |  |  |  |  |  |  |
| **CCI 2-4** |  |  |  |  |  |  |  |  |
| No Booster | 128,034 | 10,594,310 | 93 | 0.9<br>(0.7-1.1) | - | 0.9 | - |  |
| Booster | 129,993 | 10,847,400 | 24 | 0.2<br>(0.2-0.4) | -0.7<br>(-0.9--0.5) | 0.2 | 76.2<br>(62.5-84.9) |  |
| **CCI ≥ 5** |  |  |  |  |  |  |  |  |
| No Booster | 28,030 | 2,258,123 | 79 | 3.9<br>(3.0-4.9) | - | 3.5 | - |  |
| Booster | 28,126 | 2,293,908 | 13 | 0.7<br>(0.4-1.2) | -3.2<br>(-4.1--2.3) | 0.6 | 84.2<br>(70.6-91.5) |  |

\* Only matched sets still at risk 10 days after time zero. Characteristics of these participants are shown in Supplementary Table 5
\*\* Indicates one or both of the groups has cell counts less than 10.

### Negative outcome control

Cumulative incidence of SARS-CoV-2 infection during the first 10 days after time zero was nearly identical in the booster and matched no booster cohorts (**Figure 1d** and **Table 2**), suggesting adequate matching and lack of substantial unmeasured confounding.

## Discussion

Our target trial emulation study performed in the national VA healthcare system demonstrated that mRNA booster vaccination administered at least 5 months after primary vaccination had an estimated VE of 42.3% (40.6-43.9) against infection, 53.3% (48.1-58.0) against hospitalization and 79.1% (71.2-84.9) against death during follow-up extending up to 121 days in an omicron variant predominant era. Booster VE was similar for different booster types (BNT162b2 or mRNA-1273), age groups and primary vaccination regimens, and increased with longer time since primary vaccination and with greater comorbidity burden. These findings provide strong support for current CDC recommendations that all persons should receive a booster vaccination >5 months after completion of primary immunization.

Booster VE against SARS-CoV-2 infection with the delta variant was found to be very high in a phase 3 randomized controlled trial^4^ and observational cohort studies from Israel^5, 6^, ranging from 86% to 95.3%. However, a population-based matched cohort study from Qatar reported that mRNA booster VE in the omicron era was much lower at 49.4% (47.1-51.6) against symptomatic infection and 76.5% (55.9-87.5) against hospitalization or death^13^, with follow-up extending up to 35 days. Unlike the population we studied, only 9% of the population of Qatar are over age 50, few were reported to have serious coexisting conditions and no individual-level comorbid conditions were available for matching, adjustment or subgroup analysis. A target trial emulation using matched cohorts derived from nationwide population registries in Spain estimated that mRNA booster VE against infection in the omicron era was 51.3% (50.2-52.4) with follow-up extending only up to 34 days^14^ – comorbid conditions were unavailable for matching, adjustment or sub-group analyses. A test-negative, case-control study from England reported that booster VE against infection was 67.2% (66.5-67.8) at 2-4 weeks after receipt of a BNT162b2 booster before declining to 45.7% (44.7-46.7) at 10 or more weeks^15^. We found even lower booster VE in the omicron era against infection (42.3% [40.6-43.9]) and hospitalization (53.3% [48.1-58.0]), which may be related to the greater comorbidity burden of our study population and the longer follow-up extending up to 121 days. Taken together these studies suggest that booster VE against infection and hospitalization is lower in the omicron era than it was in the delta era and likely declines over time.

Our study demonstrated a much higher booster VE against SARS-CoV-2-related death (79.1% [71.2-84.9]), which was not assessed in prior studies in the omicron era. An Israeli population-based study conducted in the delta era reported that BNT162b2 booster recipients had 90% lower mortality due to Covid-19 than participants who did not receive a booster^7^, which is higher than what we found in the omicron era.

Some studies reported that mRNA-1273 booster VE against infection was slightly higher than BNT162b2 booster VE^14^ but others did not^13^. We found that booster VE against infection, hospitalization and death was not significantly different between mRNA-1273 and BNT162b2. Future target trials specifically designed to compare mRNA-1273 versus BNT162b2 boosters are needed to address this question directly.

As in prior studies^13, 14^, we found that booster VE against infection and hospitalization was significantly higher as more time accrued since primary vaccination (>9 vs. 5-9 months). This is most likely related to the waning protection of primary vaccination over time in the comparator, no-booster group. Booster VE was also substantially higher in groups with greater comorbidity burden. Since these groups also have the highest absolute risk of adverse COVID-19 outcomes, this resulted in much greater risk differences in infection, hospitalization and death between the booster and no booster groups in persons with high comorbidity burden. These results reinforce the need to ensure booster vaccination especially in persons with high comorbidity burden.

Receipt of booster dose vaccination has been documented in only 35.8% of the United States population as of 06/12/2022, a far lower proportion than countries such as Uruguay (76.1.3%), Iceland (67.9%), Italy (67.5%), Germany (58.7%), the United Kingdom (58.3%) and Israel (57.2%)^2^. Our data suggest that achieving higher rates of booster vaccination represents an important opportunity to reduce SARS-CoV-2 related morbidity and mortality. At the same time the lower booster VE against omicron than was previously observed against delta variants suggests that development of new generation of broadly effective vaccines, including potentially “universal coronavirus vaccines”, may be a better strategy than repeated booster vaccination with existing vaccines^38^.

Our study has the following limitations. Despite adherence to principles of target trial emulation, a matching method that resulted in balanced distribution of baseline characteristics in the two arms and adjustment for potential confounders, residual confounding cannot be completely excluded in a non-randomized study. However, our analysis of a negative outcome control (infection rate in the first 10 days after time zero) suggested little confounding. While some additional infections (diagnosed or undiagnosed) and hospitalizations undoubtedly occurred and were not captured in our analysis, we would expect this outcome misclassification to be nondifferential. Non-differential misclassification would have minimal impact on relative measures of effect (i.e. VE), although absolute risks and risk differences may have been slightly underestimated. If any booster doses were missed (i.e. not administered within VA or VA community care or documented in VA records or Medicare data) among matched comparators, such misclassification would tend to attenuate the reported VE. Our study population is predominantly male, which may limit the generalizability of our findings to women.

In conclusion, in a target trial emulation study of older Veterans with high burden of comorbidity conducted in the omicron era, mRNA booster VE against SARS-CoV-2 infection (42.3%) and SARS-CoV-2-related hospitalization (53.3%) were slightly lower than previously reported but VE against SARS-CoV-2-related death was high (79.1%). Improved booster vaccination should be pursued to reduce COVID-19 morbidity and mortality especially in persons with multiple comorbid conditions.

## Data Availability

Data produced in the present study are available only to investigators who have appropriate regulatory approval according to the regulations of the Veterans Affairs Office of Research and Development

## Authors’ Contributions and Authorship Statement

Drs. Kristin Berry and George Ioannou and had full access to all the data in the study and take responsibility for the integrity of the data and the accuracy of the data analysis

Dr. George Ioannou is the guarantor of this paper. All authors approved the final version of the manuscript.

Ioannou, G: Study concept and design, interpretation of results, drafting of manuscript, critical revision of manuscript, obtaining funding.

Bohnert: Study design, Interpretation of results, critical revision of manuscript, obtaining funding

O’Hare: critical revision of manuscript, obtaining funding.

Boyko: Study design, critical revision of manuscript, obtaining funding.

Maciejewski: Study design, critical revision of manuscript, obtaining funding.

Smith: Study design, critical revision of manuscript, obtaining funding.

Bowling: critical revision of manuscript, obtaining funding.

Viglianti: critical revision of manuscript, obtaining funding.

Iwashyna: critical revision of manuscript, obtaining funding.

Hynes: critical revision of manuscript, obtaining funding.

Berry: Study concept and design, statistical analysis, interpretation of results, drafting of manuscript, critical revision of manuscript.

Green: Obtaining access to data, creation of analytic variables and datasets

Fox: Obtaining access to data, creation of analytic variables and datasets

Korpak: Obtaining access to data, creation of analytic variables and datasets

Shahoumian: Obtaining access to data, creation of analytic variables and datasets

Hickock: Obtaining access to data, creation of analytic variables and datasets

Rowneki: Obtaining access to data, creation of analytic variables and datasets

Wang: Obtaining access to data, creation of analytic variables and datasets

Locke: Obtaining access to data, creation of analytic variables and datasets

## Declaration of Personal Interests

None of the authors has any conflicts of interest to disclose.

## Supplementary Appendix

**Supplementary Table 1.**
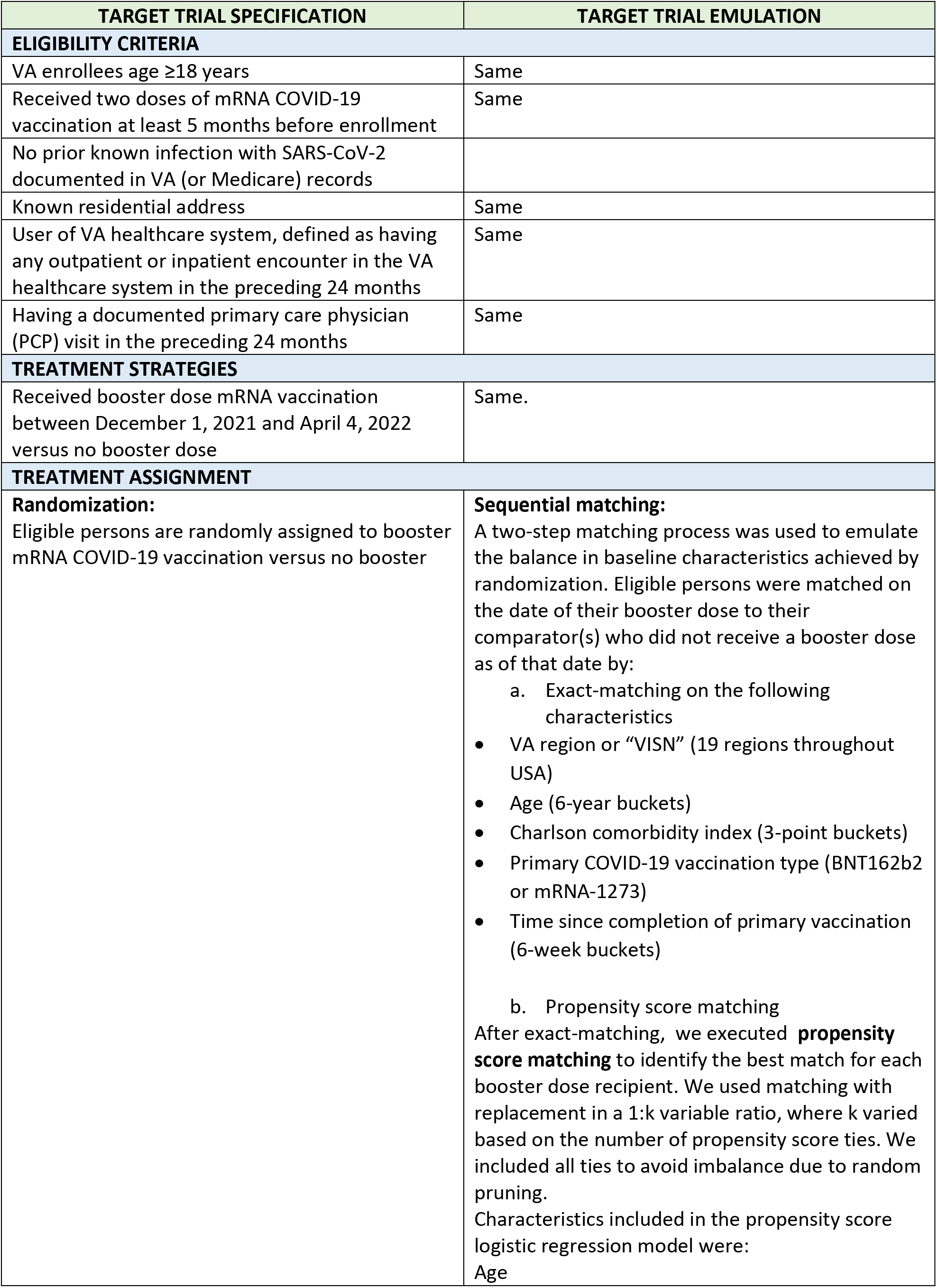
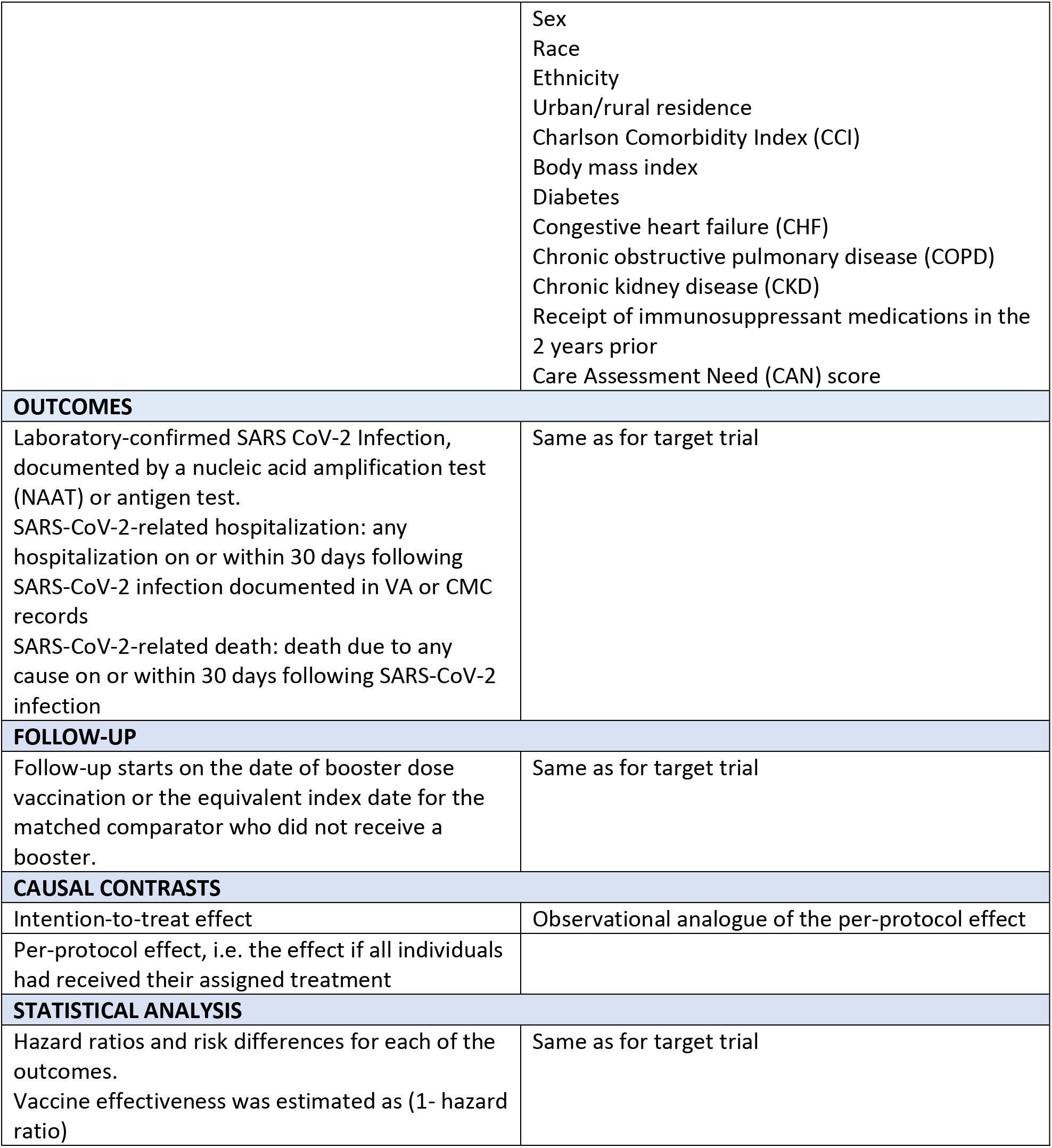
Specification and emulation of a target trial evaluating the effectiveness of booster (i.e. third dose) mRNA COVID-19 vaccination conducted from December 1, 2021 to March 31, 2022.

| TARGET TRIAL SPECIFICATION | TARGET TRIAL EMULATION |
| --- | --- |
| **ELIGIBILITY CRITERIA** |  |
| VA enrollees age $\geq 18$ years | Same |
| Received two doses of mRNA COVID-19 vaccination at least 5 months before enrollment | Same |
| No prior known infection with SARS-CoV-2 documented in VA (or Medicare) records |  |
| Known residential address | Same |
| User of VA healthcare system, defined as having any outpatient or inpatient encounter in the VA healthcare system in the preceding 24 months | Same |
| Having a documented primary care physician (PCP) visit in the preceding 24 months | Same |
| **TREATMENT STRATEGIES** |  |
| Received booster dose mRNA vaccination between December 1, 2021 and April 4, 2022 versus no booster dose | Same. |
| **TREATMENT ASSIGNMENT** |  |
| **Randomization:**<br>Eligible persons are randomly assigned to booster mRNA COVID-19 vaccination versus no booster | **Sequential matching:**<br>A two-step matching process was used to emulate the balance in baseline characteristics achieved by randomization. Eligible persons were matched on the date of their booster dose to their comparator(s) who did not receive a booster dose as of that date by:<br>a. Exact-matching on the following characteristics<br>• VA region or “VISN” (19 regions throughout USA)<br>• Age (6-year buckets)<br>• Charlson comorbidity index (3-point buckets)<br>• Primary COVID-19 vaccination type (BNT162b2 or mRNA-1273)<br>• Time since completion of primary vaccination (6-week buckets)<br>b. Propensity score matching<br>After exact-matching, we executed **propensity score matching** to identify the best match for each booster dose recipient. We used matching with replacement in a 1:k variable ratio, where k varied based on the number of propensity score ties. We included all ties to avoid imbalance due to random pruning.<br>Characteristics included in the propensity score logistic regression model were:<br>Age |
|  | Sex<br>Race<br>Ethnicity<br>Urban/rural residence<br>Charlson Comorbidity Index (CCI)<br>Body mass index<br>Diabetes<br>Congestive heart failure (CHF)<br>Chronic obstructive pulmonary disease (COPD)<br>Chronic kidney disease (CKD)<br>Receipt of immunosuppressant medications in the 2 years prior<br>Care Assessment Need (CAN) score |
| **OUTCOMES** |  |
| Laboratory-confirmed SARS CoV-2 Infection, documented by a nucleic acid amplification test (NAAT) or antigen test.<br>SARS-CoV-2-related hospitalization: any hospitalization on or within 30 days following SARS-CoV-2 infection documented in VA or CMC records<br>SARS-CoV-2-related death: death due to any cause on or within 30 days following SARS-CoV-2 infection | Same as for target trial |
| **FOLLOW-UP** |  |
| Follow-up starts on the date of booster dose vaccination or the equivalent index date for the matched comparator who did not receive a booster. | Same as for target trial |
| **CAUSAL CONTRASTS** |  |
| Intention-to-treat effect | Observational analogue of the per-protocol effect |
| Per-protocol effect, i.e. the effect if all individuals had received their assigned treatment |  |
| **STATISTICAL ANALYSIS** |  |
| Hazard ratios and risk differences for each of the outcomes.<br>Vaccine effectiveness was estimated as (1- hazard ratio) | Same as for target trial |

**Supplementary Table 2.** Immunosuppressive and cancer medications. We identified prescription of these medications within the 2-year period prior to the study period as evidence of being on potentially immune suppressive medications

| Drug class | Drugs |
| --- | --- |
| Alkylating agents | bendamustine, busulfan, chlorambucil, cisplatin, cyclophosphamide, ifosfamide, lurbinectedin, melphalan, nitrogen mustard procarbazine, temozolomide |
| Anti-CD19/CD3 | blinatumomab |
| Anti-HER2 | trastuzumab |
| Antimetabolites | azathioprine, capecitabine, cladribine, cytarabine, fludarabine, floxuridine, fluorouracil, gemcitabine, mercaptopurine, methotrexate, pemetrexed, pentostatin, pralatrexate, thioguanine |
| Antitumor antibiotics | bleomycin, dacarbazine, dactinomycin, daunorubicin, doxorubicin, epirubicin, idarubicin, mitomycin, mitoxantrone |
| B-cell depletion and inhibition | belimumab, ibriumomab, tiuxetan, obinutuzumab, ocrelizumab,<sup>b</sup> ofatumumab,<sup>b</sup> rituximab<sup>b</sup> |
| BCL2 inhibitors | venetoxlax |
| Interferon | interferon alpha-2a, interferon alpha-2b, interferon beta-1a, interferon beta-1b, interferon gamma, interferon gamma-1b |
| BCR-ABL tyrosine kinase inhibitors | bosutinib, dasatinib, imatinib, nilotinib, ponatinib |
| Bruton tyrosine kinase (BTK) inhibitors | acalabrutinib, ibrutinib, zanubrutinib |
| Calcineurin inhibitors | cyclosporine, pimecrolimus, tacrolimus, voclosporin |
| Complement inhibitor | eculizumab |
| EGFR inhibitors | afatinib, cetuximab, dacomitinib, erlotinib, gefitinib, lapatinib, mobocertinib, necitumumab, osimertinib, panitumumab |
| Glucocorticoids (systemic) | cortisone acetate, betamethasone, budesonide, dexamethasone, hydrocortisone, methylprednisolone, prednisolone, prednisone |
| Histone deacetylase inhibitors | belinostat, vorinostat |
| IL-1 inhibitors | anakinra, canakinumab, rilonacept |
| IL-12/23 inhibitors | guselkumab, ustekinumab |
| IL-17 inhibitors | brodalumab, ixekizumab, secukinumab |
| IL-4/13 inhibitors | dupilumab |
| IL-5 inhibitors | benralizumab, mepolizumab, reslizumab |
| IL-6 inhibitors | sarilumab, siltuximab, tocilizumab |
| Immune checkpoint inhibitors (anti-CTLA-4) | ipilimumab |
| Immune checkpoint inhibitors (anti-PD-1) | cemiplimab, dostarlimab, nivolumab, pembrolizumab |
| Immune checkpoint inhibitors (anti-PD-L1) | atezolizumab, avelumab, durvalumab |
| Immune globulin | anti-thymocyte globulin<sup>b</sup> |
| Immunosuppressant agent | mycophenolate mofetil (MMF), mycophenolate sodium |
| Janus kinase (JAK) inhibitors | baricitinib, ruxolitinib, tofacitinib, upadacitinib |
| Miscellaneous antineoplastics | arsenic trioxide, blinhydroxyurea, idelalisib, olaparib, omacetaxine, palbociclib, pegaspargase, thalidomide, trabectedin |
| Miscellaneous biological | glatiramer acetate |
| Miscellaneous other | leflunomide, sulfasalazine |
| mTOR kinase inhibitors | everolimus, sirolimus, temsirolimus |
| Nitrosoureas | carmustine, lomustine, streptozocin |
| Platinum derivatives | carboplatin, oxaliplatin |
| Proteasome inhibitors | bortezomib, carfilzomib |
| Selective adhesion-molecule inhibitors | natalizumab, vedolizumab |
| T-cell costimulation inhibitors | abatacept, belatacept, alemtuzumab,<sup>2</sup> basiliximab |
| Thalidomide analogs | lenalidomide, pomalidomide |
| TNF inhibitors | adalimumab, certolizumab, etanercept, golimumab, infliximab |
| Topoisomerase inhibitors | irinotecan, mitoxantrone, topotecan |
| Tubulin-acting agents | eribulin, etoposide, ixabepilone |
| Tubulin-acting agents (taxanes) | cabazitaxel, docetaxel, nabpaclitaxel, paclitaxel |
| Tubulin-acting agents (vinca alkaloids) | vinblastine, vincristine, vinorelbine |
| VEGF tyrosine kinase inhibitors | aflibercept, axitinib, bevacizumab, lenvatinib, pazopanib, ramucirumab, regorafenib, sorafenib, sunitinib, vandetanib |

**Supplementary Table 3:** ICD10 Codes used to define diabetes, congestive heart failure (CHF), chronic obstructive pulmonary disease (COPD) and chronic kidney disease (CKD). The presence of these conditions was defined by documentation of one or more of these ICD10 codes recorded in VA electronic health records (both outpatient or inpatient records) during the 2-year period prior to the date of vaccination. We used lists of ICD10 codes developed by the VA’s Centralized Interactive Phenomics Resource or CIPHER to define each condition. CIPHER develops and validates EHR-based phenotype algorithms and definitions. CIPHER is available through VA intranet at: https://vhacdwdwhweb100.vha.med.va.gov/phenotype/index.php/VA_Phenomics_Library_-_Centralized_Interactive_Phenomics_Resource_(CIPHER)

| Phenotype | Code | CodeDescription |
| --- | --- | --- |
| **CONGESTIVE HEART FAILURE (CHF)** |  |  |
| CHF | I50.20 | Unspecified systolic (congestive) heart failure |
| CHF | I50.21 | Acute systolic (congestive) heart failure |
| CHF | I50.22 | Chronic systolic (congestive) heart failure |
| CHF | I50.23 | Acute on chronic systolic (congestive) heart failure |
| CHF | I50.30 | Unspecified diastolic (congestive) heart failure |
| CHF | I50.31 | Acute diastolic (congestive) heart failure |
| CHF | I50.32 | Chronic diastolic (congestive) heart failure |
| CHF | I50.33 | Acute on chronic diastolic (congestive) heart failure |
| CHF | I50.40 | Unspecified combined systolic (congestive) and diastolic (congestive) heart failure |
| CHF | I50.41 | Acute combined systolic (congestive) and diastolic (congestive) heart failure |
| **CHRONIC KIDNEY DISEASE (CKD) AND KIDNEY FAILURE (CKF)** |  |  |
| CKD | 585.9 | CHRONIC KIDNEY DISEASE, UNSPECIFIED |
| CKD | D63.1 | Anemia in chronic kidney disease |
| CKD | E08.22 | Diabetes mellitus due to underlying condition with diabetic chronic kidney disease |
| CKD | E09.22 | Drug or chemical induced diabetes mellitus with diabetic chronic kidney disease |
| CKD | E10.22 | Type 1 diabetes mellitus with diabetic chronic kidney disease |
| CKD | E11.22 | Type 2 diabetes mellitus with diabetic chronic kidney disease |
| CKD | E13.22 | Other specified diabetes mellitus with diabetic chronic kidney disease |
| CKD | I12.0 | Hypertensive chronic kidney disease with stage 5 chronic kidney disease or end stage renal disease |
| CKD | I12.9 | Hypertensive chronic kidney disease with stage 1 through stage 4 chronic kidney disease, or unspecified chronic kidney disease |
| CKD | I13.0 | Hypertensive heart and chronic kidney disease with heart failure and stage 1 through stage 4 chronic kidney disease, or unspecified chronic kidney disease |
| CKD | I13.10 | Hypertensive heart and chronic kidney disease without heart failure, with stage 1 through stage 4 chronic kidney disease, or unspecified chronic kidney disease |
| CKD | I13.11 | Hypertensive heart and chronic kidney disease without heart failure, with stage 5 chronic kidney disease, or end stage renal disease |
| CKD | I13.2 | Hypertensive heart and chronic kidney disease with heart failure and with stage 5 chronic kidney disease, or end stage renal disease |
| CKD | N18.1 | Chronic kidney disease, stage 1 |
| CKD | N18.2 | Chronic kidney disease, stage 2 (mild) |
| CKD | N18.3 | Chronic kidney disease, stage 3 (moderate) |
| CKD | N18.30 | Chronic kidney disease, stage 3 unspecified |
| CKD | N18.31 | Chronic kidney disease, stage 3a |
| CKD | N18.32 | Chronic kidney disease, stage 3b |
| CKD | N18.4 | Chronic kidney disease, stage 4 (severe) |
| CKD | N18.5 | Chronic kidney disease, stage 5 |
| CKD | N18.6 | End stage renal disease |
| CKD | N18.9 | Chronic kidney disease, unspecified |
| CKD | R88.0 | Cloudy (hemodialysis) (peritoneal) dialysis effluent |
| CKD | Z49.01 | Encounter for fitting and adjustment of extracorporeal dialysis catheter |
| CKD | Z49.02 | Encounter for fitting and adjustment of peritoneal dialysis catheter |
| CKD | Z49.31 | Encounter for adequacy testing for hemodialysis |
| CKD | Z49.32 | Encounter for adequacy testing for peritoneal dialysis |
| CKF | I12.0 | Hypertensive chronic kidney disease with stage 5 chronic kidney disease or end stage renal disease |
| CKF | I13.11 | Hypertensive heart and chronic kidney disease without heart failure, with stage 5 chronic kidney disease, or end stage renal disease |
| CKF | I13.2 | Hypertensive heart and chronic kidney disease with heart failure and with stage 5 chronic kidney disease, or end stage renal disease |
| CKF | N18.6 | End stage renal disease |
| CKF | N99.0 | Postprocedural (acute) (chronic) kidney failure |
| **CHRONIC OBSTRUCTIVE PULMONARY DISEASE (COPD)** |  |  |
| COPD | J40. | Bronchitis, not specified as acute or chronic |
| COPD | J41.0 | Simple chronic bronchitis |
| COPD | J41.1 | Mucopurulent chronic bronchitis |
| COPD | J41.8 | Mixed simple and mucopurulent chronic bronchitis |
| COPD | J42. | Unspecified chronic bronchitis |
| COPD | J43.0 | Unilateral pulmonary emphysema [MacLeod's syndrome] |
| COPD | J43.1 | Panlobular emphysema |
| COPD | J43.2 | Centrilobular emphysema |
| COPD | J43.8 | Other emphysema |
| COPD | J43.9 | Emphysema, unspecified |
| COPD | J44.0 | Chronic obstructive pulmonary disease with (acute) lower respiratory infection |
| COPD | J44.1 | Chronic obstructive pulmonary disease with (acute) exacerbation |
| COPD | J44.9 | Chronic obstructive pulmonary disease, unspecified |
| COPD | J47.0 | Bronchiectasis with acute lower respiratory infection |
| COPD | J47.1 | Bronchiectasis with (acute) exacerbation |
| COPD | J47.9 | Bronchiectasis, uncomplicated |
| **DIABETES** |  |  |
| DiabetesAny | 250.00 | DIABETES MELLITUS WITHOUT MENTION OF COMPLICATION, TYPE II OR UNSPECIFIED TYPE, NOT STATED AS UNCONTROLLED |
| DiabetesAny | 250.60 | DIABETES WITH NEUROLOGICAL MANIFESTATIONS, TYPE II OR UNSPECIFIED TYPE, NOT STATED AS UNCONTROLLED |
| DiabetesAny | E08.00 | Diabetes mellitus due to underlying condition with hyperosmolarity without nonketotic hyperglycemic-hyperosmolar coma (NKHHC) |
| DiabetesAny | E08.01 | Diabetes mellitus due to underlying condition with hyperosmolarity with coma |
| DiabetesAny | E08.10 | Diabetes mellitus due to underlying condition with ketoacidosis without coma |
| DiabetesAny | E08.11 | Diabetes mellitus due to underlying condition with ketoacidosis with coma |
| DiabetesAny | E08.21 | Diabetes mellitus due to underlying condition with diabetic nephropathy |
| DiabetesAny | E08.22 | Diabetes mellitus due to underlying condition with diabetic chronic kidney disease |
| DiabetesAny | E08.29 | Diabetes mellitus due to underlying condition with other diabetic kidney complication |
| DiabetesAny | E08.311 | Diabetes mellitus due to underlying condition with unspecified diabetic retinopathy with macular edema |
| DiabetesAny | E08.319 | Diabetes mellitus due to underlying condition with unspecified diabetic retinopathy without macular edema |
| DiabetesAny | E08.3211 | Diabetes mellitus due to underlying condition with mild nonproliferative diabetic retinopathy with macular edema, right eye |
| DiabetesAny | E08.3212 | Diabetes mellitus due to underlying condition with mild nonproliferative diabetic retinopathy with macular edema, left eye |
| DiabetesAny | E08.3213 | Diabetes mellitus due to underlying condition with mild nonproliferative diabetic retinopathy with macular edema, bilateral |
| DiabetesAny | E08.3219 | Diabetes mellitus due to underlying condition with mild nonproliferative diabetic retinopathy with macular edema, unspecified eye |
| DiabetesAny | E08.3291 | Diabetes mellitus due to underlying condition with mild nonproliferative diabetic retinopathy without macular edema, right eye |
| DiabetesAny | E08.3292 | Diabetes mellitus due to underlying condition with mild nonproliferative diabetic retinopathy without macular edema, left eye |
| DiabetesAny | E08.3293 | Diabetes mellitus due to underlying condition with mild nonproliferative diabetic retinopathy without macular edema, bilateral |
| DiabetesAny | E08.3299 | Diabetes mellitus due to underlying condition with mild nonproliferative diabetic retinopathy without macular edema, unspecified eye |
| DiabetesAny | E08.3311 | Diabetes mellitus due to underlying condition with moderate nonproliferative diabetic retinopathy with macular edema, right eye |
| DiabetesAny | E08.3312 | Diabetes mellitus due to underlying condition with moderate nonproliferative diabetic retinopathy with macular edema, left eye |
| DiabetesAny | E08.3313 | Diabetes mellitus due to underlying condition with moderate nonproliferative diabetic retinopathy with macular edema, bilateral |
| DiabetesAny | E08.3319 | Diabetes mellitus due to underlying condition with moderate nonproliferative diabetic retinopathy with macular edema, unspecified eye |
| DiabetesAny | E08.3391 | Diabetes mellitus due to underlying condition with moderate nonproliferative diabetic retinopathy without macular edema, right eye |
| DiabetesAny | E08.3392 | Diabetes mellitus due to underlying condition with moderate nonproliferative diabetic retinopathy without macular edema, left eye |
| DiabetesAny | E08.3393 | Diabetes mellitus due to underlying condition with moderate nonproliferative diabetic retinopathy without macular edema, bilateral |
| DiabetesAny | E08.3399 | Diabetes mellitus due to underlying condition with moderate nonproliferative diabetic retinopathy without macular edema, unspecified eye |
| DiabetesAny | E08.3411 | Diabetes mellitus due to underlying condition with severe nonproliferative diabetic retinopathy with macular edema, right eye |
| DiabetesAny | E08.3412 | Diabetes mellitus due to underlying condition with severe nonproliferative diabetic retinopathy with macular edema, left eye |
| DiabetesAny | E08.3413 | Diabetes mellitus due to underlying condition with severe nonproliferative diabetic retinopathy with macular edema, bilateral |
| DiabetesAny | E08.3419 | Diabetes mellitus due to underlying condition with severe nonproliferative diabetic retinopathy with macular edema, unspecified eye |
| DiabetesAny | E08.3491 | Diabetes mellitus due to underlying condition with severe nonproliferative diabetic retinopathy without macular edema, right eye |
| DiabetesAny | E08.3492 | Diabetes mellitus due to underlying condition with severe nonproliferative diabetic retinopathy without macular edema, left eye |
| DiabetesAny | E08.3493 | Diabetes mellitus due to underlying condition with severe nonproliferative diabetic retinopathy without macular edema, bilateral |
| DiabetesAny | E08.3499 | Diabetes mellitus due to underlying condition with severe nonproliferative diabetic retinopathy without macular edema, unspecified eye |
| DiabetesAny | E08.3511 | Diabetes mellitus due to underlying condition with proliferative diabetic retinopathy with macular edema, right eye |
| DiabetesAny | E08.3512 | Diabetes mellitus due to underlying condition with proliferative diabetic retinopathy with macular edema, left eye |
| DiabetesAny | E08.3513 | Diabetes mellitus due to underlying condition with proliferative diabetic retinopathy with macular edema, bilateral |
| DiabetesAny | E08.3519 | Diabetes mellitus due to underlying condition with proliferative diabetic retinopathy with macular edema, unspecified eye |
| DiabetesAny | E08.3521 | Diabetes mellitus due to underlying condition with proliferative diabetic retinopathy with traction retinal detachment involving the macula, right eye |
| DiabetesAny | E08.3522 | Diabetes mellitus due to underlying condition with proliferative diabetic retinopathy with traction retinal detachment involving the macula, left eye |
| DiabetesAny | E08.3523 | Diabetes mellitus due to underlying condition with proliferative diabetic retinopathy with traction retinal detachment involving the macula, bilateral |
| DiabetesAny | E08.3529 | Diabetes mellitus due to underlying condition with proliferative diabetic retinopathy with traction retinal detachment involving the macula, unspecified eye |
| DiabetesAny | E08.3531 | Diabetes mellitus due to underlying condition with proliferative diabetic retinopathy with traction retinal detachment not involving the macula, right eye |
| DiabetesAny | E08.3532 | Diabetes mellitus due to underlying condition with proliferative diabetic retinopathy with traction retinal detachment not involving the macula, left eye |
| DiabetesAny | E08.3533 | Diabetes mellitus due to underlying condition with proliferative diabetic retinopathy with traction retinal detachment not involving the macula, bilateral |
| DiabetesAny | E08.3539 | Diabetes mellitus due to underlying condition with proliferative diabetic retinopathy with traction retinal detachment not involving the macula, unspecified eye |
| DiabetesAny | E08.3541 | Diabetes mellitus due to underlying condition with proliferative diabetic retinopathy with combined traction retinal detachment and rhegmatogenous retinal detachment, right eye |
| DiabetesAny | E08.3542 | Diabetes mellitus due to underlying condition with proliferative diabetic retinopathy with combined traction retinal detachment and rhegmatogenous retinal detachment, left eye |
| DiabetesAny | E08.3543 | Diabetes mellitus due to underlying condition with proliferative diabetic retinopathy with combined traction retinal detachment and rhegmatogenous retinal detachment, bilateral |
| DiabetesAny | E08.3549 | Diabetes mellitus due to underlying condition with proliferative diabetic retinopathy with combined traction retinal detachment and rhegmatogenous retinal detachment, unspecified |
| DiabetesAny | E08.3551 | Diabetes mellitus due to underlying condition with stable proliferative diabetic retinopathy, right eye |
| DiabetesAny | E08.3552 | Diabetes mellitus due to underlying condition with stable proliferative diabetic retinopathy, left eye |
| DiabetesAny | E08.3553 | Diabetes mellitus due to underlying condition with stable proliferative diabetic retinopathy, bilateral |
| DiabetesAny | E08.3559 | Diabetes mellitus due to underlying condition with stable proliferative diabetic retinopathy, unspecified eye |
| DiabetesAny | E08.3591 | Diabetes mellitus due to underlying condition with proliferative diabetic retinopathy without macular edema, right eye |
| DiabetesAny | E08.3592 | Diabetes mellitus due to underlying condition with proliferative diabetic retinopathy without macular edema, left eye |
| DiabetesAny | E08.3593 | Diabetes mellitus due to underlying condition with proliferative diabetic retinopathy without macular edema, bilateral |
| DiabetesAny | E08.3599 | Diabetes mellitus due to underlying condition with proliferative diabetic retinopathy without macular edema, unspecified eye |
| DiabetesAny | E08.36 | Diabetes mellitus due to underlying condition with diabetic cataract |
| DiabetesAny | E08.37X1 | Diabetes mellitus due to underlying condition with diabetic macular edema, resolved following treatment, right eye |
| DiabetesAny | E08.37X2 | Diabetes mellitus due to underlying condition with diabetic macular edema, resolved following treatment, left eye |
| DiabetesAny | E08.37X3 | Diabetes mellitus due to underlying condition with diabetic macular edema, resolved following treatment, bilateral |
| DiabetesAny | E08.37X9 | Diabetes mellitus due to underlying condition with diabetic macular edema, resolved following treatment, unspecified eye |
| DiabetesAny | E08.39 | Diabetes mellitus due to underlying condition with other diabetic ophthalmic complication |
| DiabetesAny | E08.40 | Diabetes mellitus due to underlying condition with diabetic neuropathy, unspecified |
| DiabetesAny | E08.41 | Diabetes mellitus due to underlying condition with diabetic mononeuropathy |
| DiabetesAny | E08.42 | Diabetes mellitus due to underlying condition with diabetic polyneuropathy |
| DiabetesAny | E08.43 | Diabetes mellitus due to underlying condition with diabetic autonomic (poly)neuropathy |
| DiabetesAny | E08.44 | Diabetes mellitus due to underlying condition with diabetic amyotrophy |
| DiabetesAny | E08.49 | Diabetes mellitus due to underlying condition with other diabetic neurological complication |
| DiabetesAny | E08.51 | Diabetes mellitus due to underlying condition with diabetic peripheral angiopathy without gangrene |
| DiabetesAny | E08.52 | Diabetes mellitus due to underlying condition with diabetic peripheral angiopathy with gangrene |
| DiabetesAny | E08.59 | Diabetes mellitus due to underlying condition with other circulatory complications |
| DiabetesAny | E08.610 | Diabetes mellitus due to underlying condition with diabetic neuropathic arthropathy |
| DiabetesAny | E08.618 | Diabetes mellitus due to underlying condition with other diabetic arthropathy |
| DiabetesAny | E08.620 | Diabetes mellitus due to underlying condition with diabetic dermatitis |
| DiabetesAny | E08.621 | Diabetes mellitus due to underlying condition with foot ulcer |
| DiabetesAny | E08.622 | Diabetes mellitus due to underlying condition with other skin ulcer |
| DiabetesAny | E08.628 | Diabetes mellitus due to underlying condition with other skin complications |
| DiabetesAny | E08.630 | Diabetes mellitus due to underlying condition with periodontal disease |
| DiabetesAny | E08.638 | Diabetes mellitus due to underlying condition with other oral complications |
| DiabetesAny | E08.641 | Diabetes mellitus due to underlying condition with hypoglycemia with coma |
| DiabetesAny | E08.649 | Diabetes mellitus due to underlying condition with hypoglycemia without coma |
| DiabetesAny | E08.65 | Diabetes mellitus due to underlying condition with hyperglycemia |
| DiabetesAny | E08.69 | Diabetes mellitus due to underlying condition with other specified complication |
| DiabetesAny | E08.8 | Diabetes mellitus due to underlying condition with unspecified complications |
| DiabetesAny | E08.9 | Diabetes mellitus due to underlying condition without complications |
| DiabetesAny | E09.00 | Drug or chemical induced diabetes mellitus with hyperosmolarity without nonketotic hyperglycemic-hyperosmolar coma (NKHHC) |
| DiabetesAny | E09.01 | Drug or chemical induced diabetes mellitus with hyperosmolarity with coma |
| DiabetesAny | E09.10 | Drug or chemical induced diabetes mellitus with ketoacidosis without coma |
| DiabetesAny | E09.11 | Drug or chemical induced diabetes mellitus with ketoacidosis with coma |
| DiabetesAny | E09.21 | Drug or chemical induced diabetes mellitus with diabetic nephropathy |
| DiabetesAny | E09.22 | Drug or chemical induced diabetes mellitus with diabetic chronic kidney disease |
| DiabetesAny | E09.29 | Drug or chemical induced diabetes mellitus with other diabetic kidney complication |
| DiabetesAny | E09.311 | Drug or chemical induced diabetes mellitus with unspecified diabetic retinopathy with macular edema |
| DiabetesAny | E09.319 | Drug or chemical induced diabetes mellitus with unspecified diabetic retinopathy without macular edema |
| DiabetesAny | E09.3211 | Drug or chemical induced diabetes mellitus with mild nonproliferative diabetic retinopathy with macular edema, right eye |
| DiabetesAny | E09.3212 | Drug or chemical induced diabetes mellitus with mild nonproliferative diabetic retinopathy with macular edema, left eye |
| DiabetesAny | E09.3213 | Drug or chemical induced diabetes mellitus with mild nonproliferative diabetic retinopathy with macular edema, bilateral |
| DiabetesAny | E09.3219 | Drug or chemical induced diabetes mellitus with mild nonproliferative diabetic retinopathy with macular edema, unspecified eye |
| DiabetesAny | E09.3291 | Drug or chemical induced diabetes mellitus with mild nonproliferative diabetic retinopathy without macular edema, right eye |
| DiabetesAny | E09.3292 | Drug or chemical induced diabetes mellitus with mild nonproliferative diabetic retinopathy without macular edema, left eye |
| DiabetesAny | E09.3293 | Drug or chemical induced diabetes mellitus with mild nonproliferative diabetic retinopathy without macular edema, bilateral |
| DiabetesAny | E09.3299 | Drug or chemical induced diabetes mellitus with mild nonproliferative diabetic retinopathy without macular edema, unspecified eye |
| DiabetesAny | E09.3311 | Drug or chemical induced diabetes mellitus with moderate nonproliferative diabetic retinopathy with macular edema, right eye |
| DiabetesAny | E09.3312 | Drug or chemical induced diabetes mellitus with moderate nonproliferative diabetic retinopathy with macular edema, left eye |
| DiabetesAny | E09.3313 | Drug or chemical induced diabetes mellitus with moderate nonproliferative diabetic retinopathy with macular edema, bilateral |
| DiabetesAny | E09.3319 | Drug or chemical induced diabetes mellitus with moderate nonproliferative diabetic retinopathy with macular edema, unspecified eye |
| DiabetesAny | E09.3391 | Drug or chemical induced diabetes mellitus with moderate nonproliferative diabetic retinopathy without macular edema, right eye |
| DiabetesAny | E09.3392 | Drug or chemical induced diabetes mellitus with moderate nonproliferative diabetic retinopathy without macular edema, left eye |
| DiabetesAny | E09.3393 | Drug or chemical induced diabetes mellitus with moderate nonproliferative diabetic retinopathy without macular edema, bilateral |
| DiabetesAny | E09.3399 | Drug or chemical induced diabetes mellitus with moderate nonproliferative diabetic retinopathy without macular edema, unspecified eye |
| DiabetesAny | E09.3411 | Drug or chemical induced diabetes mellitus with severe nonproliferative diabetic retinopathy with macular edema, right eye |
| DiabetesAny | E09.3412 | Drug or chemical induced diabetes mellitus with severe nonproliferative diabetic retinopathy with macular edema, left eye |
| DiabetesAny | E09.3413 | Drug or chemical induced diabetes mellitus with severe nonproliferative diabetic retinopathy with macular edema, bilateral |
| DiabetesAny | E09.3419 | Drug or chemical induced diabetes mellitus with severe nonproliferative diabetic retinopathy with macular edema, unspecified eye |
| DiabetesAny | E09.3493 | Drug or chemical induced diabetes mellitus with severe nonproliferative diabetic retinopathy without macular edema, bilateral |
| DiabetesAny | E09.3499 | Drug or chemical induced diabetes mellitus with severe nonproliferative diabetic retinopathy without macular edema, unspecified eye |
| DiabetesAny | E09.3511 | Drug or chemical induced diabetes mellitus with proliferative diabetic retinopathy with macular edema, right eye |
| DiabetesAny | E09.3512 | Drug or chemical induced diabetes mellitus with proliferative diabetic retinopathy with macular edema, left eye |
| DiabetesAny | E09.3513 | Drug or chemical induced diabetes mellitus with proliferative diabetic retinopathy with macular edema, bilateral |
| DiabetesAny | E09.3519 | Drug or chemical induced diabetes mellitus with proliferative diabetic retinopathy with macular edema, unspecified eye |
| DiabetesAny | E09.3522 | Drug or chemical induced diabetes mellitus with proliferative diabetic retinopathy with traction retinal detachment involving the macula, left eye |
| DiabetesAny | E09.3533 | Drug or chemical induced diabetes mellitus with proliferative diabetic retinopathy with traction retinal detachment not involving the macula, bilateral |
| DiabetesAny | E09.3541 | Drug or chemical induced diabetes mellitus with proliferative diabetic retinopathy with combined traction retinal detachment and rhegmatogenous retinal detachment, right eye |
| DiabetesAny | E09.3542 | Drug or chemical induced diabetes mellitus with proliferative diabetic retinopathy with combined traction retinal detachment and rhegmatogenous retinal detachment, left eye |
| DiabetesAny | E09.3551 | Drug or chemical induced diabetes mellitus with stable proliferative diabetic retinopathy, right eye |
| DiabetesAny | E09.3552 | Drug or chemical induced diabetes mellitus with stable proliferative diabetic retinopathy, left eye |
| DiabetesAny | E09.3553 | Drug or chemical induced diabetes mellitus with stable proliferative diabetic retinopathy, bilateral |
| DiabetesAny | E09.3559 | Drug or chemical induced diabetes mellitus with stable proliferative diabetic retinopathy, unspecified eye |
| DiabetesAny | E09.3591 | Drug or chemical induced diabetes mellitus with proliferative diabetic retinopathy without macular edema, right eye |
| DiabetesAny | E09.3592 | Drug or chemical induced diabetes mellitus with proliferative diabetic retinopathy without macular edema, left eye |
| DiabetesAny | E09.3593 | Drug or chemical induced diabetes mellitus with proliferative diabetic retinopathy without macular edema, bilateral |
| DiabetesAny | E09.3599 | Drug or chemical induced diabetes mellitus with proliferative diabetic retinopathy without macular edema, unspecified eye |
| DiabetesAny | E09.36 | Drug or chemical induced diabetes mellitus with diabetic cataract |
| DiabetesAny | E09.37X1 | Drug or chemical induced diabetes mellitus with diabetic macular edema, resolved following treatment, right eye |
| DiabetesAny | E09.37X2 | Drug or chemical induced diabetes mellitus with diabetic macular edema, resolved following treatment, left eye |
| DiabetesAny | E09.37X3 | Drug or chemical induced diabetes mellitus with diabetic macular edema, resolved following treatment, bilateral |
| DiabetesAny | E09.39 | Drug or chemical induced diabetes mellitus with other diabetic ophthalmic complication |
| DiabetesAny | E09.40 | Drug or chemical induced diabetes mellitus with neurological complications with diabetic neuropathy, unspecified |
| DiabetesAny | E09.41 | Drug or chemical induced diabetes mellitus with neurological complications with diabetic mononeuropathy |
| DiabetesAny | E09.42 | Drug or chemical induced diabetes mellitus with neurological complications with diabetic polyneuropathy |
| DiabetesAny | E09.43 | Drug or chemical induced diabetes mellitus with neurological complications with diabetic autonomic (poly)neuropathy |
| DiabetesAny | E09.44 | Drug or chemical induced diabetes mellitus with neurological complications with diabetic amyotrophy |
| DiabetesAny | E09.49 | Drug or chemical induced diabetes mellitus with neurological complications with other diabetic neurological complication |
| DiabetesAny | E09.51 | Drug or chemical induced diabetes mellitus with diabetic peripheral angiopathy without gangrene |
| DiabetesAny | E09.52 | Drug or chemical induced diabetes mellitus with diabetic peripheral angiopathy with gangrene |
| DiabetesAny | E09.59 | Drug or chemical induced diabetes mellitus with other circulatory complications |
| DiabetesAny | E09.610 | Drug or chemical induced diabetes mellitus with diabetic neuropathic arthropathy |
| DiabetesAny | E09.618 | Drug or chemical induced diabetes mellitus with other diabetic arthropathy |
| DiabetesAny | E09.620 | Drug or chemical induced diabetes mellitus with diabetic dermatitis |
| DiabetesAny | E09.621 | Drug or chemical induced diabetes mellitus with foot ulcer |
| DiabetesAny | E09.622 | Drug or chemical induced diabetes mellitus with other skin ulcer |
| DiabetesAny | E09.628 | Drug or chemical induced diabetes mellitus with other skin complications |
| DiabetesAny | E09.630 | Drug or chemical induced diabetes mellitus with periodontal disease |
| DiabetesAny | E09.638 | Drug or chemical induced diabetes mellitus with other oral complications |
| DiabetesAny | E09.641 | Drug or chemical induced diabetes mellitus with hypoglycemia with coma |
| DiabetesAny | E09.649 | Drug or chemical induced diabetes mellitus with hypoglycemia without coma |
| DiabetesAny | E09.65 | Drug or chemical induced diabetes mellitus with hyperglycemia |
| DiabetesAny | E09.69 | Drug or chemical induced diabetes mellitus with other specified complication |
| DiabetesAny | E09.8 | Drug or chemical induced diabetes mellitus with unspecified complications |
| DiabetesAny | E09.9 | Drug or chemical induced diabetes mellitus without complications |
| DiabetesAny | E10.10 | Type 1 diabetes mellitus with ketoacidosis without coma |
| DiabetesAny | E10.11 | Type 1 diabetes mellitus with ketoacidosis with coma |
| DiabetesAny | E10.21 | Type 1 diabetes mellitus with diabetic nephropathy |
| DiabetesAny | E10.22 | Type 1 diabetes mellitus with diabetic chronic kidney disease |
| DiabetesAny | E10.29 | Type 1 diabetes mellitus with other diabetic kidney complication |
| DiabetesAny | E10.311 | Type 1 diabetes mellitus with unspecified diabetic retinopathy with macular edema |
| DiabetesAny | E10.319 | Type 1 diabetes mellitus with unspecified diabetic retinopathy without macular edema |
| DiabetesAny | E10.3211 | Type 1 diabetes mellitus with mild nonproliferative diabetic retinopathy with macular edema, right eye |
| DiabetesAny | E10.3212 | Type 1 diabetes mellitus with mild nonproliferative diabetic retinopathy with macular edema, left eye |
| DiabetesAny | E10.3213 | Type 1 diabetes mellitus with mild nonproliferative diabetic retinopathy with macular edema, bilateral |
| DiabetesAny | E10.3219 | Type 1 diabetes mellitus with mild nonproliferative diabetic retinopathy with macular edema, unspecified eye |
| DiabetesAny | E10.3291 | Type 1 diabetes mellitus with mild nonproliferative diabetic retinopathy without macular edema, right eye |
| DiabetesAny | E10.3292 | Type 1 diabetes mellitus with mild nonproliferative diabetic retinopathy without macular edema, left eye |
| DiabetesAny | E10.3293 | Type 1 diabetes mellitus with mild nonproliferative diabetic retinopathy without macular edema, bilateral |
| DiabetesAny | E10.3299 | Type 1 diabetes mellitus with mild nonproliferative diabetic retinopathy without macular edema, unspecified eye |
| DiabetesAny | E10.3311 | Type 1 diabetes mellitus with moderate nonproliferative diabetic retinopathy with macular edema, right eye |
| DiabetesAny | E10.3312 | Type 1 diabetes mellitus with moderate nonproliferative diabetic retinopathy with macular edema, left eye |
| DiabetesAny | E10.3313 | Type 1 diabetes mellitus with moderate nonproliferative diabetic retinopathy with macular edema, bilateral |
| DiabetesAny | E10.3319 | Type 1 diabetes mellitus with moderate nonproliferative diabetic retinopathy with macular edema, unspecified eye |
| DiabetesAny | E10.3391 | Type 1 diabetes mellitus with moderate nonproliferative diabetic retinopathy without macular edema, right eye |
| DiabetesAny | E10.3392 | Type 1 diabetes mellitus with moderate nonproliferative diabetic retinopathy without macular edema, left eye |
| DiabetesAny | E10.3393 | Type 1 diabetes mellitus with moderate nonproliferative diabetic retinopathy without macular edema, bilateral |
| DiabetesAny | E10.3399 | Type 1 diabetes mellitus with moderate nonproliferative diabetic retinopathy without macular edema, unspecified eye |
| DiabetesAny | E10.3411 | Type 1 diabetes mellitus with severe nonproliferative diabetic retinopathy with macular edema, right eye |
| DiabetesAny | E10.3412 | Type 1 diabetes mellitus with severe nonproliferative diabetic retinopathy with macular edema, left eye |
| DiabetesAny | E10.3413 | Type 1 diabetes mellitus with severe nonproliferative diabetic retinopathy with macular edema, bilateral |
| DiabetesAny | E10.3419 | Type 1 diabetes mellitus with severe nonproliferative diabetic retinopathy with macular edema, unspecified eye |
| DiabetesAny | E10.3491 | Type 1 diabetes mellitus with severe nonproliferative diabetic retinopathy without macular edema, right eye |
| DiabetesAny | E10.3492 | Type 1 diabetes mellitus with severe nonproliferative diabetic retinopathy without macular edema, left eye |
| DiabetesAny | E10.3493 | Type 1 diabetes mellitus with severe nonproliferative diabetic retinopathy without macular edema, bilateral |
| DiabetesAny | E10.3499 | Type 1 diabetes mellitus with severe nonproliferative diabetic retinopathy without macular edema, unspecified eye |
| DiabetesAny | E10.3511 | Type 1 diabetes mellitus with proliferative diabetic retinopathy with macular edema, right eye |
| DiabetesAny | E10.3512 | Type 1 diabetes mellitus with proliferative diabetic retinopathy with macular edema, left eye |
| DiabetesAny | E10.3513 | Type 1 diabetes mellitus with proliferative diabetic retinopathy with macular edema, bilateral |
| DiabetesAny | E10.3519 | Type 1 diabetes mellitus with proliferative diabetic retinopathy with macular edema, unspecified eye |
| DiabetesAny | E10.3521 | Type 1 diabetes mellitus with proliferative diabetic retinopathy with traction retinal detachment involving the macula, right eye |
| DiabetesAny | E10.3522 | Type 1 diabetes mellitus with proliferative diabetic retinopathy with traction retinal detachment involving the macula, left eye |
| DiabetesAny | E10.3523 | Type 1 diabetes mellitus with proliferative diabetic retinopathy with traction retinal detachment involving the macula, bilateral |
| DiabetesAny | E10.3529 | Type 1 diabetes mellitus with proliferative diabetic retinopathy with traction retinal detachment involving the macula, unspecified eye |
| DiabetesAny | E10.3531 | Type 1 diabetes mellitus with proliferative diabetic retinopathy with traction retinal detachment not involving the macula, right eye |
| DiabetesAny | E10.3532 | Type 1 diabetes mellitus with proliferative diabetic retinopathy with traction retinal detachment not involving the macula, left eye |
| DiabetesAny | E10.3533 | Type 1 diabetes mellitus with proliferative diabetic retinopathy with traction retinal detachment not involving the macula, bilateral |
| DiabetesAny | E10.3539 | Type 1 diabetes mellitus with proliferative diabetic retinopathy with traction retinal detachment not involving the macula, unspecified eye |
| DiabetesAny | E10.3541 | Type 1 diabetes mellitus with proliferative diabetic retinopathy with combined traction retinal detachment and rhegmatogenous retinal detachment, right eye |
| DiabetesAny | E10.3542 | Type 1 diabetes mellitus with proliferative diabetic retinopathy with combined traction retinal detachment and rhegmatogenous retinal detachment, left eye |
| DiabetesAny | E10.3543 | Type 1 diabetes mellitus with proliferative diabetic retinopathy with combined traction retinal detachment and rhegmatogenous retinal detachment, bilateral |
| DiabetesAny | E10.3549 | Type 1 diabetes mellitus with proliferative diabetic retinopathy with combined traction retinal detachment and rhegmatogenous retinal detachment, unspecified eye |
| DiabetesAny | E10.3551 | Type 1 diabetes mellitus with stable proliferative diabetic retinopathy, right eye |
| DiabetesAny | E10.3552 | Type 1 diabetes mellitus with stable proliferative diabetic retinopathy, left eye |
| DiabetesAny | E10.3553 | Type 1 diabetes mellitus with stable proliferative diabetic retinopathy, bilateral |
| DiabetesAny | E10.3559 | Type 1 diabetes mellitus with stable proliferative diabetic retinopathy, unspecified eye |
| DiabetesAny | E10.3591 | Type 1 diabetes mellitus with proliferative diabetic retinopathy without macular edema, right eye |
| DiabetesAny | E10.3592 | Type 1 diabetes mellitus with proliferative diabetic retinopathy without macular edema, left eye |
| DiabetesAny | E10.3593 | Type 1 diabetes mellitus with proliferative diabetic retinopathy without macular edema, bilateral |
| DiabetesAny | E10.3599 | Type 1 diabetes mellitus with proliferative diabetic retinopathy without macular edema, unspecified eye |
| DiabetesAny | E10.36 | Type 1 diabetes mellitus with diabetic cataract |
| DiabetesAny | E10.37X1 | Type 1 diabetes mellitus with diabetic macular edema, resolved following treatment, right eye |
| DiabetesAny | E10.37X2 | Type 1 diabetes mellitus with diabetic macular edema, resolved following treatment, left eye |
| DiabetesAny | E10.37X3 | Type 1 diabetes mellitus with diabetic macular edema, resolved following treatment, bilateral |
| DiabetesAny | E10.37X9 | Type 1 diabetes mellitus with diabetic macular edema, resolved following treatment, unspecified eye |
| DiabetesAny | E10.39 | Type 1 diabetes mellitus with other diabetic ophthalmic complication |
| DiabetesAny | E10.40 | Type 1 diabetes mellitus with diabetic neuropathy, unspecified |
| DiabetesAny | E10.41 | Type 1 diabetes mellitus with diabetic mononeuropathy |
| DiabetesAny | E10.42 | Type 1 diabetes mellitus with diabetic polyneuropathy |
| DiabetesAny | E10.43 | Type 1 diabetes mellitus with diabetic autonomic (poly)neuropathy |
| DiabetesAny | E10.44 | Type 1 diabetes mellitus with diabetic amyotrophy |
| DiabetesAny | E10.49 | Type 1 diabetes mellitus with other diabetic neurological complication |
| DiabetesAny | E10.51 | Type 1 diabetes mellitus with diabetic peripheral angiopathy without gangrene |
| DiabetesAny | E10.52 | Type 1 diabetes mellitus with diabetic peripheral angiopathy with gangrene |
| DiabetesAny | E10.59 | Type 1 diabetes mellitus with other circulatory complications |
| DiabetesAny | E10.610 | Type 1 diabetes mellitus with diabetic neuropathic arthropathy |
| DiabetesAny | E10.618 | Type 1 diabetes mellitus with other diabetic arthropathy |
| DiabetesAny | E10.620 | Type 1 diabetes mellitus with diabetic dermatitis |
| DiabetesAny | E10.621 | Type 1 diabetes mellitus with foot ulcer |
| DiabetesAny | E10.622 | Type 1 diabetes mellitus with other skin ulcer |
| DiabetesAny | E10.628 | Type 1 diabetes mellitus with other skin complications |
| DiabetesAny | E10.630 | Type 1 diabetes mellitus with periodontal disease |
| DiabetesAny | E10.638 | Type 1 diabetes mellitus with other oral complications |
| DiabetesAny | E10.641 | Type 1 diabetes mellitus with hypoglycemia with coma |
| DiabetesAny | E10.649 | Type 1 diabetes mellitus with hypoglycemia without coma |
| DiabetesAny | E10.65 | Type 1 diabetes mellitus with hyperglycemia |
| DiabetesAny | E10.69 | Type 1 diabetes mellitus with other specified complication |
| DiabetesAny | E10.8 | Type 1 diabetes mellitus with unspecified complications |
| DiabetesAny | E10.9 | Type 1 diabetes mellitus without complications |
| DiabetesAny | E11.00 | Type 2 diabetes mellitus with hyperosmolarity without nonketotic hyperglycemic-hyperosmolar coma (NKHHC) |
| DiabetesAny | E11.01 | Type 2 diabetes mellitus with hyperosmolarity with coma |
| DiabetesAny | E11.10 | Type 2 diabetes mellitus with ketoacidosis without coma |
| DiabetesAny | E11.11 | Type 2 diabetes mellitus with ketoacidosis with coma |
| DiabetesAny | E11.21 | Type 2 diabetes mellitus with diabetic nephropathy |
| DiabetesAny | E11.22 | Type 2 diabetes mellitus with diabetic chronic kidney disease |
| DiabetesAny | E11.29 | Type 2 diabetes mellitus with other diabetic kidney complication |
| DiabetesAny | E11.311 | Type 2 diabetes mellitus with unspecified diabetic retinopathy with macular edema |
| DiabetesAny | E11.319 | Type 2 diabetes mellitus with unspecified diabetic retinopathy without macular edema |
| DiabetesAny | E11.321 | Type 2 Diabetes Mellitus with Mild Nonproliferative Diabetic Retinopathy with Macular Edema |
| DiabetesAny | E11.3211 | Type 2 diabetes mellitus with mild nonproliferative diabetic retinopathy with macular edema, right eye |
| DiabetesAny | E11.3212 | Type 2 diabetes mellitus with mild nonproliferative diabetic retinopathy with macular edema, left eye |
| DiabetesAny | E11.3213 | Type 2 diabetes mellitus with mild nonproliferative diabetic retinopathy with macular edema, bilateral |
| DiabetesAny | E11.3219 | Type 2 diabetes mellitus with mild nonproliferative diabetic retinopathy with macular edema, unspecified eye |
| DiabetesAny | E11.329 | Type 2 Diabetes Mellitus with Mild Nonproliferative Diabetic Retinopathy without Macular Edema |
| DiabetesAny | E11.3291 | Type 2 diabetes mellitus with mild nonproliferative diabetic retinopathy without macular edema, right eye |
| DiabetesAny | E11.3292 | Type 2 diabetes mellitus with mild nonproliferative diabetic retinopathy without macular edema, left eye |
| DiabetesAny | E11.3293 | Type 2 diabetes mellitus with mild nonproliferative diabetic retinopathy without macular edema, bilateral |
| DiabetesAny | E11.3299 | Type 2 diabetes mellitus with mild nonproliferative diabetic retinopathy without macular edema, unspecified eye |
| DiabetesAny | E11.3311 | Type 2 diabetes mellitus with moderate nonproliferative diabetic retinopathy with macular edema, right eye |
| DiabetesAny | E11.3312 | Type 2 diabetes mellitus with moderate nonproliferative diabetic retinopathy with macular edema, left eye |
| DiabetesAny | E11.3313 | Type 2 diabetes mellitus with moderate nonproliferative diabetic retinopathy with macular edema, bilateral |
| DiabetesAny | E11.3319 | Type 2 diabetes mellitus with moderate nonproliferative diabetic retinopathy with macular edema, unspecified eye |
| DiabetesAny | E11.339 | Type 2 Diabetes Mellitus with Moderate Nonproliferative Diabetic Retinopathy without Macular Edema |
| DiabetesAny | E11.3391 | Type 2 diabetes mellitus with moderate nonproliferative diabetic retinopathy without macular edema, right eye |
| DiabetesAny | E11.3392 | Type 2 diabetes mellitus with moderate nonproliferative diabetic retinopathy without macular edema, left eye |
| DiabetesAny | E11.3393 | Type 2 diabetes mellitus with moderate nonproliferative diabetic retinopathy without macular edema, bilateral |
| DiabetesAny | E11.3399 | Type 2 diabetes mellitus with moderate nonproliferative diabetic retinopathy without macular edema, unspecified eye |
| DiabetesAny | E11.341 | Type 2 Diabetes Mellitus with Severe Nonproliferative Diabetic Retinopathy with Macular Edema |
| DiabetesAny | E11.3411 | Type 2 diabetes mellitus with severe nonproliferative diabetic retinopathy with macular edema, right eye |
| DiabetesAny | E11.3412 | Type 2 diabetes mellitus with severe nonproliferative diabetic retinopathy with macular edema, left eye |
| DiabetesAny | E11.3413 | Type 2 diabetes mellitus with severe nonproliferative diabetic retinopathy with macular edema, bilateral |
| DiabetesAny | E11.3419 | Type 2 diabetes mellitus with severe nonproliferative diabetic retinopathy with macular edema, unspecified eye |
| DiabetesAny | E11.3491 | Type 2 diabetes mellitus with severe nonproliferative diabetic retinopathy without macular edema, right eye |
| DiabetesAny | E11.3492 | Type 2 diabetes mellitus with severe nonproliferative diabetic retinopathy without macular edema, left eye |
| DiabetesAny | E11.3493 | Type 2 diabetes mellitus with severe nonproliferative diabetic retinopathy without macular edema, bilateral |
| DiabetesAny | E11.3499 | Type 2 diabetes mellitus with severe nonproliferative diabetic retinopathy without macular edema, unspecified eye |
| DiabetesAny | E11.351 | Type 2 Diabetes Mellitus with Proliferative Diabetic Retinopathy with Macular Edema |
| DiabetesAny | E11.3511 | Type 2 diabetes mellitus with proliferative diabetic retinopathy with macular edema, right eye |
| DiabetesAny | E11.3512 | Type 2 diabetes mellitus with proliferative diabetic retinopathy with macular edema, left eye |
| DiabetesAny | E11.3513 | Type 2 diabetes mellitus with proliferative diabetic retinopathy with macular edema, bilateral |
| DiabetesAny | E11.3519 | Type 2 diabetes mellitus with proliferative diabetic retinopathy with macular edema, unspecified eye |
| DiabetesAny | E11.3521 | Type 2 diabetes mellitus with proliferative diabetic retinopathy with traction retinal detachment involving the macula, right eye |
| DiabetesAny | E11.3522 | Type 2 diabetes mellitus with proliferative diabetic retinopathy with traction retinal detachment involving the macula, left eye |
| DiabetesAny | E11.3523 | Type 2 diabetes mellitus with proliferative diabetic retinopathy with traction retinal detachment involving the macula, bilateral |
| DiabetesAny | E11.3529 | Type 2 diabetes mellitus with proliferative diabetic retinopathy with traction retinal detachment involving the macula, unspecified eye |
| DiabetesAny | E11.3531 | Type 2 diabetes mellitus with proliferative diabetic retinopathy with traction retinal detachment not involving the macula, right eye |
| DiabetesAny | E11.3532 | Type 2 diabetes mellitus with proliferative diabetic retinopathy with traction retinal detachment not involving the macula, left eye |
| DiabetesAny | E11.3533 | Type 2 diabetes mellitus with proliferative diabetic retinopathy with traction retinal detachment not involving the macula, bilateral |
| DiabetesAny | E11.3539 | Type 2 diabetes mellitus with proliferative diabetic retinopathy with traction retinal detachment not involving the macula, unspecified eye |
| DiabetesAny | E11.3541 | Type 2 diabetes mellitus with proliferative diabetic retinopathy with combined traction retinal detachment and rhegmatogenous retinal detachment, right eye |
| DiabetesAny | E11.3542 | Type 2 diabetes mellitus with proliferative diabetic retinopathy with combined traction retinal detachment and rhegmatogenous retinal detachment, left eye |
| DiabetesAny | E11.3543 | Type 2 diabetes mellitus with proliferative diabetic retinopathy with combined traction retinal detachment and rhegmatogenous retinal detachment, bilateral |
| DiabetesAny | E11.3549 | Type 2 diabetes mellitus with proliferative diabetic retinopathy with combined traction retinal detachment and rhegmatogenous retinal detachment, unspecified eye |
| DiabetesAny | E11.3551 | Type 2 diabetes mellitus with stable proliferative diabetic retinopathy, right eye |
| DiabetesAny | E11.3552 | Type 2 diabetes mellitus with stable proliferative diabetic retinopathy, left eye |
| DiabetesAny | E11.3553 | Type 2 diabetes mellitus with stable proliferative diabetic retinopathy, bilateral |
| DiabetesAny | E11.3559 | Type 2 diabetes mellitus with stable proliferative diabetic retinopathy, unspecified eye |
| DiabetesAny | E11.359 | Type 2 Diabetes Mellitus with Proliferative Diabetic Retinopathy without Macular Edema |
| DiabetesAny | E11.3591 | Type 2 diabetes mellitus with proliferative diabetic retinopathy without macular edema, right eye |
| DiabetesAny | E11.3592 | Type 2 diabetes mellitus with proliferative diabetic retinopathy without macular edema, left eye |
| DiabetesAny | E11.3593 | Type 2 diabetes mellitus with proliferative diabetic retinopathy without macular edema, bilateral |
| DiabetesAny | E11.3599 | Type 2 diabetes mellitus with proliferative diabetic retinopathy without macular edema, unspecified eye |
| DiabetesAny | E11.36 | Type 2 diabetes mellitus with diabetic cataract |
| DiabetesAny | E11.37X1 | Type 2 diabetes mellitus with diabetic macular edema, resolved following treatment, right eye |
| DiabetesAny | E11.37X2 | Type 2 diabetes mellitus with diabetic macular edema, resolved following treatment, left eye |
| DiabetesAny | E11.37X3 | Type 2 diabetes mellitus with diabetic macular edema, resolved following treatment, bilateral |
| DiabetesAny | E11.37X9 | Type 2 diabetes mellitus with diabetic macular edema, resolved following treatment, unspecified eye |
| DiabetesAny | E11.39 | Type 2 diabetes mellitus with other diabetic ophthalmic complication |
| DiabetesAny | E11.40 | Type 2 diabetes mellitus with diabetic neuropathy, unspecified |
| DiabetesAny | E11.41 | Type 2 diabetes mellitus with diabetic mononeuropathy |
| DiabetesAny | E11.42 | Type 2 diabetes mellitus with diabetic polyneuropathy |
| DiabetesAny | E11.43 | Type 2 diabetes mellitus with diabetic autonomic (poly)neuropathy |
| DiabetesAny | E11.44 | Type 2 diabetes mellitus with diabetic amyotrophy |
| DiabetesAny | E11.49 | Type 2 diabetes mellitus with other diabetic neurological complication |
| DiabetesAny | E11.51 | Type 2 diabetes mellitus with diabetic peripheral angiopathy without gangrene |
| DiabetesAny | E11.52 | Type 2 diabetes mellitus with diabetic peripheral angiopathy with gangrene |
| DiabetesAny | E11.59 | Type 2 diabetes mellitus with other circulatory complications |
| DiabetesAny | E11.610 | Type 2 diabetes mellitus with diabetic neuropathic arthropathy |
| DiabetesAny | E11.618 | Type 2 diabetes mellitus with other diabetic arthropathy |
| DiabetesAny | E11.620 | Type 2 diabetes mellitus with diabetic dermatitis |
| DiabetesAny | E11.621 | Type 2 diabetes mellitus with foot ulcer |
| DiabetesAny | E11.622 | Type 2 diabetes mellitus with other skin ulcer |
| DiabetesAny | E11.628 | Type 2 diabetes mellitus with other skin complications |
| DiabetesAny | E11.630 | Type 2 diabetes mellitus with periodontal disease |
| DiabetesAny | E11.638 | Type 2 diabetes mellitus with other oral complications |
| DiabetesAny | E11.641 | Type 2 diabetes mellitus with hypoglycemia with coma |
| DiabetesAny | E11.649 | Type 2 diabetes mellitus with hypoglycemia without coma |
| DiabetesAny | E11.65 | Type 2 diabetes mellitus with hyperglycemia |
| DiabetesAny | E11.69 | Type 2 diabetes mellitus with other specified complication |
| DiabetesAny | E11.8 | Type 2 diabetes mellitus with unspecified complications |
| DiabetesAny | E11.9 | Type 2 diabetes mellitus without complications |
| DiabetesAny | E13.00 | Other specified diabetes mellitus with hyperosmolarity without nonketotic hyperglycemic-hyperosmolar coma (NKHHC) |
| DiabetesAny | E13.01 | Other specified diabetes mellitus with hyperosmolarity with coma |
| DiabetesAny | E13.10 | Other specified diabetes mellitus with ketoacidosis without coma |
| DiabetesAny | E13.11 | Other specified diabetes mellitus with ketoacidosis with coma |
| DiabetesAny | E13.21 | Other specified diabetes mellitus with diabetic nephropathy |
| DiabetesAny | E13.22 | Other specified diabetes mellitus with diabetic chronic kidney disease |
| DiabetesAny | E13.29 | Other specified diabetes mellitus with other diabetic kidney complication |
| DiabetesAny | E13.311 | Other specified diabetes mellitus with unspecified diabetic retinopathy with macular edema |
| DiabetesAny | E13.319 | Other specified diabetes mellitus with unspecified diabetic retinopathy without macular edema |
| DiabetesAny | E13.3211 | Other specified diabetes mellitus with mild nonproliferative diabetic retinopathy with macular edema, right eye |
| DiabetesAny | E13.3212 | Other specified diabetes mellitus with mild nonproliferative diabetic retinopathy with macular edema, left eye |
| DiabetesAny | E13.3213 | Other specified diabetes mellitus with mild nonproliferative diabetic retinopathy with macular edema, bilateral |
| DiabetesAny | E13.3219 | Other specified diabetes mellitus with mild nonproliferative diabetic retinopathy with macular edema, unspecified eye |
| DiabetesAny | E13.3291 | Other specified diabetes mellitus with mild nonproliferative diabetic retinopathy without macular edema, right eye |
| DiabetesAny | E13.3292 | Other specified diabetes mellitus with mild nonproliferative diabetic retinopathy without macular edema, left eye |
| DiabetesAny | E13.3293 | Other specified diabetes mellitus with mild nonproliferative diabetic retinopathy without macular edema, bilateral |
| DiabetesAny | E13.3299 | Other specified diabetes mellitus with mild nonproliferative diabetic retinopathy without macular edema, unspecified eye |
| DiabetesAny | E13.3311 | Other specified diabetes mellitus with moderate nonproliferative diabetic retinopathy with macular edema, right eye |
| DiabetesAny | E13.3312 | Other specified diabetes mellitus with moderate nonproliferative diabetic retinopathy with macular edema, left eye |
| DiabetesAny | E13.3313 | Other specified diabetes mellitus with moderate nonproliferative diabetic retinopathy with macular edema, bilateral |
| DiabetesAny | E13.3319 | Other specified diabetes mellitus with moderate nonproliferative diabetic retinopathy with macular edema, unspecified eye |
| DiabetesAny | E13.3391 | Other specified diabetes mellitus with moderate nonproliferative diabetic retinopathy without macular edema, right eye |
| DiabetesAny | E13.3392 | Other specified diabetes mellitus with moderate nonproliferative diabetic retinopathy without macular edema, left eye |
| DiabetesAny | E13.3393 | Other specified diabetes mellitus with moderate nonproliferative diabetic retinopathy without macular edema, bilateral |
| DiabetesAny | E13.3399 | Other specified diabetes mellitus with moderate nonproliferative diabetic retinopathy without macular edema, unspecified eye |
| DiabetesAny | E13.3411 | Other specified diabetes mellitus with severe nonproliferative diabetic retinopathy with macular edema, right eye |
| DiabetesAny | E13.3412 | Other specified diabetes mellitus with severe nonproliferative diabetic retinopathy with macular edema, left eye |
| DiabetesAny | E13.3413 | Other specified diabetes mellitus with severe nonproliferative diabetic retinopathy with macular edema, bilateral |
| DiabetesAny | E13.3419 | Other specified diabetes mellitus with severe nonproliferative diabetic retinopathy with macular edema, unspecified eye |
| DiabetesAny | E13.3491 | Other specified diabetes mellitus with severe nonproliferative diabetic retinopathy without macular edema, right eye |
| DiabetesAny | E13.3492 | Other specified diabetes mellitus with severe nonproliferative diabetic retinopathy without macular edema, left eye |
| DiabetesAny | E13.3493 | Other specified diabetes mellitus with severe nonproliferative diabetic retinopathy without macular edema, bilateral |
| DiabetesAny | E13.3499 | Other specified diabetes mellitus with severe nonproliferative diabetic retinopathy without macular edema, unspecified eye |
| DiabetesAny | E13.3511 | Other specified diabetes mellitus with proliferative diabetic retinopathy with macular edema, right eye |
| DiabetesAny | E13.3512 | Other specified diabetes mellitus with proliferative diabetic retinopathy with macular edema, left eye |
| DiabetesAny | E13.3513 | Other specified diabetes mellitus with proliferative diabetic retinopathy with macular edema, bilateral |
| DiabetesAny | E13.3519 | Other specified diabetes mellitus with proliferative diabetic retinopathy with macular edema, unspecified eye |
| DiabetesAny | E13.3521 | Other specified diabetes mellitus with proliferative diabetic retinopathy with traction retinal detachment involving the macula, right eye |
| DiabetesAny | E13.3522 | Other specified diabetes mellitus with proliferative diabetic retinopathy with traction retinal detachment involving the macula, left eye |
| DiabetesAny | E13.3523 | Other specified diabetes mellitus with proliferative diabetic retinopathy with traction retinal detachment involving the macula, bilateral |
| DiabetesAny | E13.3529 | Other specified diabetes mellitus with proliferative diabetic retinopathy with traction retinal detachment involving the macula, unspecified eye |
| DiabetesAny | E13.3531 | Other specified diabetes mellitus with proliferative diabetic retinopathy with traction retinal detachment not involving the macula, right eye |
| DiabetesAny | E13.3532 | Other specified diabetes mellitus with proliferative diabetic retinopathy with traction retinal detachment not involving the macula, left eye |
| DiabetesAny | E13.3533 | Other specified diabetes mellitus with proliferative diabetic retinopathy with traction retinal detachment not involving the macula, bilateral |
| DiabetesAny | E13.3539 | Other specified diabetes mellitus with proliferative diabetic retinopathy with traction retinal detachment not involving the macula, unspecified eye |
| DiabetesAny | E13.3541 | Other specified diabetes mellitus with proliferative diabetic retinopathy with combined traction retinal detachment and rhegmatogenous retinal detachment, right eye |
| DiabetesAny | E13.3542 | Other specified diabetes mellitus with proliferative diabetic retinopathy with combined traction retinal detachment and rhegmatogenous retinal detachment, left eye |
| DiabetesAny | E13.3543 | Other specified diabetes mellitus with proliferative diabetic retinopathy with combined traction retinal detachment and rhegmatogenous retinal detachment, bilateral |
| DiabetesAny | E13.3549 | Other specified diabetes mellitus with proliferative diabetic retinopathy with combined traction retinal detachment and rhegmatogenous retinal detachment, unspecified eye |
| DiabetesAny | E13.3551 | Other specified diabetes mellitus with stable proliferative diabetic retinopathy, right eye |
| DiabetesAny | E13.3552 | Other specified diabetes mellitus with stable proliferative diabetic retinopathy, left eye |
| DiabetesAny | E13.3553 | Other specified diabetes mellitus with stable proliferative diabetic retinopathy, bilateral |
| DiabetesAny | E13.3559 | Other specified diabetes mellitus with stable proliferative diabetic retinopathy, unspecified eye |
| DiabetesAny | E13.3591 | Other specified diabetes mellitus with proliferative diabetic retinopathy without macular edema, right eye |
| DiabetesAny | E13.3592 | Other specified diabetes mellitus with proliferative diabetic retinopathy without macular edema, left eye |
| DiabetesAny | E13.3593 | Other specified diabetes mellitus with proliferative diabetic retinopathy without macular edema, bilateral |
| DiabetesAny | E13.3599 | Other specified diabetes mellitus with proliferative diabetic retinopathy without macular edema, unspecified eye |
| DiabetesAny | E13.36 | Other specified diabetes mellitus with diabetic cataract |
| DiabetesAny | E13.37X1 | Other specified diabetes mellitus with diabetic macular edema, resolved following treatment, right eye |
| DiabetesAny | E13.37X2 | Other specified diabetes mellitus with diabetic macular edema, resolved following treatment, left eye |
| DiabetesAny | E13.37X3 | Other specified diabetes mellitus with diabetic macular edema, resolved following treatment, bilateral |
| DiabetesAny | E13.37X9 | Other specified diabetes mellitus with diabetic macular edema, resolved following treatment, unspecified eye |
| DiabetesAny | E13.39 | Other specified diabetes mellitus with other diabetic ophthalmic complication |
| DiabetesAny | E13.40 | Other specified diabetes mellitus with diabetic neuropathy, unspecified |
| DiabetesAny | E13.41 | Other specified diabetes mellitus with diabetic mononeuropathy |
| DiabetesAny | E13.42 | Other specified diabetes mellitus with diabetic polyneuropathy |
| DiabetesAny | E13.43 | Other specified diabetes mellitus with diabetic autonomic (poly)neuropathy |
| DiabetesAny | E13.44 | Other specified diabetes mellitus with diabetic amyotrophy |
| DiabetesAny | E13.49 | Other specified diabetes mellitus with other diabetic neurological complication |
| DiabetesAny | E13.51 | Other specified diabetes mellitus with diabetic peripheral angiopathy without gangrene |
| DiabetesAny | E13.52 | Other specified diabetes mellitus with diabetic peripheral angiopathy with gangrene |
| DiabetesAny | E13.59 | Other specified diabetes mellitus with other circulatory complications |
| DiabetesAny | E13.610 | Other specified diabetes mellitus with diabetic neuropathic arthropathy |
| DiabetesAny | E13.618 | Other specified diabetes mellitus with other diabetic arthropathy |
| DiabetesAny | E13.620 | Other specified diabetes mellitus with diabetic dermatitis |
| DiabetesAny | E13.621 | Other specified diabetes mellitus with foot ulcer |
| DiabetesAny | E13.622 | Other specified diabetes mellitus with other skin ulcer |
| DiabetesAny | E13.628 | Other specified diabetes mellitus with other skin complications |
| DiabetesAny | E13.630 | Other specified diabetes mellitus with periodontal disease |
| DiabetesAny | E13.638 | Other specified diabetes mellitus with other oral complications |
| DiabetesAny | E13.641 | Other specified diabetes mellitus with hypoglycemia with coma |
| DiabetesAny | E13.649 | Other specified diabetes mellitus with hypoglycemia without coma |
| DiabetesAny | E13.65 | Other specified diabetes mellitus with hyperglycemia |
| DiabetesAny | E13.69 | Other specified diabetes mellitus with other specified complication |
| DiabetesAny | E13.8 | Other specified diabetes mellitus with unspecified complications |
| DiabetesAny | E13.9 | Other specified diabetes mellitus without complications |
| DiabetesAny | E89.1 | Postprocedural hypoinsulinemia |
| DiabetesAny | O24.011 | Pre-existing type 1 diabetes mellitus, in pregnancy, first trimester |
| DiabetesAny | O24.012 | Pre-existing type 1 diabetes mellitus, in pregnancy, second trimester |
| DiabetesAny | O24.013 | Pre-existing type 1 diabetes mellitus, in pregnancy, third trimester |
| DiabetesAny | O24.019 | Pre-existing type 1 diabetes mellitus, in pregnancy, unspecified trimester |
| DiabetesAny | O24.02 | Pre-existing type 1 diabetes mellitus, in childbirth |
| DiabetesAny | O24.03 | Pre-existing type 1 diabetes mellitus, in the puerperium |
| DiabetesAny | O24.111 | Pre-existing type 2 diabetes mellitus, in pregnancy, first trimester |
| DiabetesAny | O24.112 | Pre-existing type 2 diabetes mellitus, in pregnancy, second trimester |
| DiabetesAny | O24.113 | Pre-existing type 2 diabetes mellitus, in pregnancy, third trimester |
| DiabetesAny | O24.119 | Pre-existing type 2 diabetes mellitus, in pregnancy, unspecified trimester |
| DiabetesAny | O24.12 | Pre-existing type 2 diabetes mellitus, in childbirth |
| DiabetesAny | O24.13 | Pre-existing type 2 diabetes mellitus, in the puerperium |
| DiabetesAny | O24.311 | Unspecified pre-existing diabetes mellitus in pregnancy, first trimester |
| DiabetesAny | O24.312 | Unspecified pre-existing diabetes mellitus in pregnancy, second trimester |
| DiabetesAny | O24.313 | Unspecified pre-existing diabetes mellitus in pregnancy, third trimester |
| DiabetesAny | O24.319 | Unspecified pre-existing diabetes mellitus in pregnancy, unspecified trimester |
| DiabetesAny | O24.32 | Unspecified pre-existing diabetes mellitus in childbirth |
| DiabetesAny | O24.33 | Unspecified pre-existing diabetes mellitus in the puerperium |
| DiabetesAny | O24.410 | Gestational diabetes mellitus in pregnancy, diet controlled |
| DiabetesAny | O24.414 | Gestational diabetes mellitus in pregnancy, insulin controlled |
| DiabetesAny | O24.415 | Gestational diabetes mellitus in pregnancy, controlled by oral hypoglycemic drugs |
| DiabetesAny | O24.419 | Gestational diabetes mellitus in pregnancy, unspecified control |
| DiabetesAny | O24.420 | Gestational diabetes mellitus in childbirth, diet controlled |
| DiabetesAny | O24.424 | Gestational diabetes mellitus in childbirth, insulin controlled |
| DiabetesAny | O24.425 | Gestational diabetes mellitus in childbirth, controlled by oral hypoglycemic drugs |
| DiabetesAny | O24.429 | Gestational diabetes mellitus in childbirth, unspecified control |
| DiabetesAny | O24.430 | Gestational diabetes mellitus in the puerperium, diet controlled |
| DiabetesAny | O24.434 | Gestational diabetes mellitus in the puerperium, insulin controlled |
| DiabetesAny | O24.435 | Gestational diabetes mellitus in puerperium, controlled by oral hypoglycemic drugs |
| DiabetesAny | O24.439 | Gestational diabetes mellitus in the puerperium, unspecified control |
| DiabetesAny | O24.811 | Other pre-existing diabetes mellitus in pregnancy, first trimester |
| DiabetesAny | O24.812 | Other pre-existing diabetes mellitus in pregnancy, second trimester |
| DiabetesAny | O24.813 | Other pre-existing diabetes mellitus in pregnancy, third trimester |
| DiabetesAny | O24.819 | Other pre-existing diabetes mellitus in pregnancy, unspecified trimester |
| DiabetesAny | O24.82 | Other pre-existing diabetes mellitus in childbirth |
| DiabetesAny | O24.83 | Other pre-existing diabetes mellitus in the puerperium |
| DiabetesAny | O24.911 | Unspecified diabetes mellitus in pregnancy, first trimester |
| DiabetesAny | O24.912 | Unspecified diabetes mellitus in pregnancy, second trimester |
| DiabetesAny | O24.913 | Unspecified diabetes mellitus in pregnancy, third trimester |
| DiabetesAny | O24.919 | Unspecified diabetes mellitus in pregnancy, unspecified trimester |
| DiabetesAny | O24.92 | Unspecified diabetes mellitus in childbirth |
| DiabetesAny | O24.93 | Unspecified diabetes mellitus in the puerperium |
| DiabetesAny | O99.810 | Abnormal glucose complicating pregnancy |
| DiabetesAny | O99.814 | Abnormal glucose complicating childbirth |
| DiabetesAny | O99.815 | Abnormal glucose complicating the puerperium |

**Supplementary Table 4.** Baseline sociodemographic and clinical characteristics of persons who received a booster dose of mRNA COVID-19 vaccination in the VA healthcare system and their unmatched counterparts who did not receive a booster dose from December 1, 2021 to March 31, 2022.

|  | **Booster mRNA**<br>**vaccination**<br>**N=505,585** | **No booster**<br>**vaccination**<br>**N=1,181,836** |
| --- | --- | --- |
| **Type of booster vaccine** |  |  |
| BNT162b2 | 37.9 | N/A |
| mRNA-1273 | 62.1 | N/A |
| **Time since 2<sup>nd</sup> dose COVID-19 mRNA vaccine (days), mean ±SD** | 274.0± 36.7 | N/A |
| **Time since 2<sup>nd</sup> dose COVID-19 mRNA vaccine (%)** |  |  |
| 5-9 months (154-270 days) | 48.0 | N/A |
| >9 months (> 270 days) | 52.0 | N/A |
| **Type of primary COVID-19 mRNA vaccine\*\*** |  |  |
| BNT162b2 | 38.5 | 43.5 |
| mRNA-1273 | 61.5 | 56.5 |
| **Sex (%)** |  |  |
| Female | 10.3 | 9.5 |
| Male | 87.3 | 87 |
| Missing | 2.3 | 3.5 |
| **Age (years), mean ±SD** | 63.0±14.0 | 62.5±16.3 |
| **Age Group (%)** |  |  |
| 18 to 49 | 17 | 22.9 |
| 50 to 59 | 19 | 15.4 |
| 60 to 64 | 14.1 | 9.7 |
| 65 to 69 | 12.1 | 10 |
| 70 to 74 | 19 | 18.6 |
| 75 to 79 | 9.7 | 10.7 |
| 80 to 84 | 4.4 | 5.5 |
| 85 to 89 | 3.2 | 4.5 |
| ≥90 | 1.6 | 2.5 |
| **Race (%)** |  |  |
| White | 65.7 | 70.8 |
| Black | 23.6 | 17.3 |
| Asian | 1.7 | 1.7 |
| American Indian/Alaska Native | 1 | 1 |
| Pacific Islander/ Native Hawaiian | 1.1 | 1 |
| Declined/Unknown/Missing | 6.9 | 8.1 |
| **Ethnicity (%)** |  |  |
| Non-Hispanic | 86.8 | 86.8 |
| Hispanic | 8.7 | 7.5 |
| Declined/Unknown/Missing | 4.4 | 5.6 |
| **Urban/Rural (%)** |  |  |
| Rural/Highly rural | 47.3 | 51.7 |
| Urban | 52.7 | 48.3 |
| **VA Integrated Service Network (VISN) (%)** |  |  |
| 1 | 4.6 | 4 |
| 2 | 5.4 | 4 |
| 4 | 5 | 4.4 |
| 5 | 2.9 | 3.4 |
| 6 | 6.5 | 6.2 |
| 7 | 6.4 | 7.9 |
| 8 | 10.3 | 9.8 |
| 9 | 3.4 | 3.8 |
| 10 | 7.7 | 6.5 |
| 12 | 5.1 | 3.5 |
| 15 | 3.3 | 3.5 |
| 16 | 6.2 | 6.1 |
| 17 | 5.3 | 7.9 |
| 19 | 3.9 | 4.9 |
| 20 | 4.1 | 6 |
| 21 | 5.8 | 4.9 |
| 22 | 9.3 | 8.6 |
| 23 | 4.9 | 4.7 |
| **Body Mass Index (kg/m<sup>2</sup>), mean ±SD** | 30.4±6.1 | 30.1±5.9 |
| **Body Mass Index (kg/m<sup>2</sup>), group (%)** |  |  |
| <18.5 | 0.7 | 0.7 |
| 18.5 to <25 | 16.4 | 16.8 |
| 25 to <30 (Overweight) | 34.1 | 34.9 |
| 30 to <35 (Obese I) | 27 | 26.1 |
| 35 to <40 (Obese II) | 12.5 | 11.5 |
| ≥40 (Obese III) | 6.7 | 5.8 |
| Missing | 2.4 | 4.3 |
| **Charlson Comorbidity Index, mean ± SD** | 1.4±1.8 | 1.1±1.6 |
| **Charlson Comorbidity Index group (%)** |  |  |
| 0 | 43.9 | 51.4 |
| 1 | 21.3 | 20.6 |
| 2 | 14.9 | 13.2 |
| 3 | 7.8 | 6.3 |
| 4 | 5.1 | 3.8 |
| 5-6 | 4.7 | 3.3 |
| 7-8 | 1.6 | 1.1 |
| ≥9 | 0.6 | 0.3 |
| **Diabetes (%)** |  |  |
| No | 68.8 | 74.2 |
| Yes | 31.2 | 25.8 |
| **Chronic Kidney Disease (%)** |  |  |
| No | 89.2 | 91.1 |
| Yes | 10.8 | 8.9 |
| **Congestive heart failure (%)** |  |  |
| No | 95.1 | 96.6 |
| Yes | 4.9 | 3.4 |
| **Chronic Obstructive Pulmonary Disease (%)** |  |  |
| No | 86.3 | 89.1 |
| Yes | 13.7 | 10.9 |
| **CAN Score† for mortality w/in 1 year, mean ± SD** | 46.8±27.8 | 46.8±28.6 |
| **CAN Score† for mortality w/in 1 year group (%)** |  |  |
| 0-30 | 33 | 34.4 |
| 31-55 | 28.4 | 24.8 |
| 56-75 | 20 | 20 |
| 76-90 | 13.2 | 14.3 |
| 95-96 | 1.7 | 1.7 |
| 97-98 | 1.6 | 1.6 |
| 99 | 0.7 | 0.6 |
| Missing | 1.4 | 2.6 |
| **Immunosuppressant medications\* (%)** |  |  |
| No | 94.1 | 95.9 |
| Yes | 5.9 | 4.1 |

† **CAN score** is the Care Assessment Needs score a validated measure of 1-year mortality in VA enrollees, presented as a percentile of all VA enrollees
\* Immunosuppressant medications prescribed in the previous year (see list of immunosuppressant medications in **Supplementary Appendix-Supplementary Methods 2**)

**Supplementary Table 5.** Baseline sociodemographic and clinical characteristics of persons who received a booster dose of mRNA COVID-19 vaccination in the VA healthcare system and their matched counterparts who did not receive a booster dose from December 1, 2021 to March 31, 2022, and for whom all members of matched sets were still at risk 10 days after time zero.

|  | **Booster**<br>**mRNA**<br>**vaccination**<br>**N=468,616** | **No booster**<br>**vaccination**<br>**N=468,616** | **Standardized**<br>**Mean**<br>**Difference** |
| --- | --- | --- | --- |
| **Type of 3<sup>rd</sup> dose COVID-19 mRNA vaccine** |  |  |  |
| BNT162b2 | 37.7 | N/A | 0.0000 |
| mRNA-1273 | 62.3 | N/A | 0.0000 |
| **Time since 2<sup>nd</sup> dose COVID-19 mRNA vaccine (days), mean ±SD** | 273.3±35.8 | 273.4±36.8 | -0.0028 |
| **Time since 2<sup>nd</sup> dose COVID-19 mRNA vaccine (%)** |  |  |  |
| 5-9 months (154-270 days) | 51.5 | 51.8 | -0.0060 |
| >9 months (> 270 days) | 48.5 | 48.2 | 0.0060 |
| **Type of primary COVID-19 mRNA vaccine\*\*** |  |  |  |
| BNT162b2 | 38.3 | 38.3 | 0.0000 |
| mRNA-1273 | 61.7 | 61.7 | 0.0000 |
| **Sex (%)** |  |  |  |
| Female | 10.2 | 9.9 | 0.0100 |
| Male | 87.4 | 87.7 | -0.0091 |
| Declined/Unknown/Missing | 2.4 | 2.4 | 0.0000 |
| **Age (years), mean ±SD** | 63.0±14.0 | 63.0±14.0 | 0.0000 |
| **Age Group (%)** |  |  |  |
| 18 to 49 | 17 | 17 | 0.0000 |
| 50 to 59 | 18.9 | 18.9 | 0.0000 |
| 60 to 64 | 14 | 13.7 | 0.0087 |
| 65 to 69 | 12.2 | 12 | 0.0061 |
| 70 to 74 | 19.1 | 19.7 | -0.0152 |
| 75 to 79 | 9.8 | 9.6 | 0.0068 |
| 80 to 84 | 4.4 | 4.4 | 0.0000 |
| 85 to 89 | 3.2 | 3.1 | 0.0057 |
| ≥90 | 1.5 | 1.5 | 0.0000 |
| **Race (%)** |  |  |  |
| White | 66.4 | 67.1 | -0.0149 |
| Black | 22.8 | 22.2 | 0.0144 |
| Asian | 1.7 | 1.7 | 0.0000 |
| American Indian/Alaska Native | 1 | 1 | 0.0000 |
| Pacific Islander/ Native Hawaiian | 1.1 | 1.1 | 0.0000 |
| Declined/Unknown/Missing | 6.9 | 6.8 | 0.0040 |
| **Ethnicity (%)** |  |  |  |
| Non-Hispanic | 86.9 | 87.3 | -0.0119 |
| Hispanic | 8.6 | 8.2 | 0.0144 |
| Declined/Unknown/Missing | 4.5 | 4.5 | 0.0000 |
| **Urban/Rural (%)** |  |  |  |
| Rural/Highly rural | 47.9 | 48.2 | -0.0060 |
| Urban | 52.1 | 51.8 | 0.0060 |
| **VA Integrated Service Network (VISN) (%)** |  |  |  |
| 1 | 4.5 | 4.5 | 0.0000 |
| 2 | 5.3 | 5.3 | 0.0000 |
| 4 | 5 | 5 | 0.0000 |
| 5 | 2.8 | 2.8 | 0.0000 |
| 6 | 6.5 | 6.5 | 0.0000 |
| 7 | 6.5 | 6.5 | 0.0000 |
| 8 | 10.4 | 10.4 | 0.0000 |
| 9 | 3.4 | 3.4 | 0.0000 |
| 10 | 7.7 | 7.7 | 0.0000 |
| 12 | 5 | 5 | 0.0000 |
| 15 | 3.2 | 3.2 | 0.0000 |
| 16 | 6.2 | 6.2 | 0.0000 |
| 17 | 5.4 | 5.4 | 0.0000 |
| 19 | 3.9 | 3.9 | 0.0000 |
| 20 | 4.1 | 4.1 | 0.0000 |
| 21 | 5.8 | 5.8 | 0.0000 |
| 22 | 9.3 | 9.3 | 0.0000 |
| 23 | 5 | 5 |  |
| **Body Mass Index (kg/m<sup>2</sup>), mean ±SD** | 30.4±6.1 | 30.4±6.1 | 0.0000 |
| **Body Mass Index (kg/m<sup>2</sup>), group (%)** |  |  |  |
| <18.5 | 0.7 | 0.7 | 0.0000 |
| 18.5 to <25 | 16.5 | 15.8 | 0.0190 |
| 25 to <30 (Overweight) | 34.2 | 33.9 | 0.0063 |
| 30 to <35 (Obese I) | 27 | 27 | 0.0000 |
| 35 to <40 (Obese II) | 12.5 | 12.6 | -0.0030 |
| ≥40 (Obese III) | 6.6 | 6.7 | -0.0040 |
| Missing | 2.5 | 3.3 | -0.0477 |
| **Charlson Comorbidity Index, mean ± SD** | 1.3±1.7 | 1.3±1.7 | 0.0000 |
| **Charlson Comorbidity Index group (%)** |  |  |  |
| 0 | 44.7 | 44.7 | 0.0000 |
| 1 | 21.5 | 22 | -0.0121 |
| 2 | 15.1 | 14.8 | 0.0084 |
| 3 | 7.8 | 7.6 | 0.0075 |
| 4 | 4.9 | 4.9 | 0.0000 |
| 5-6 | 4.4 | 4.4 | 0.0000 |
| 7-8 | 1.3 | 1.3 | 0.0000 |
| ≥9 | 0.3 | 0.3 | 0.0000 |
| **Diabetes (%)** | 30.6 | 31.2 | -0.0130 |
| **Chronic Kidney Disease (%)** | 10.2 | 10 | 0.0066 |
| **Congestive heart failure (%)** | 4.5 | 4.3 | 0.0098 |
| **Chronic Obstructive Pulmonary Disease (%)** | 13.2 | 13.5 | -0.0088 |
| **CAN Score† for mortality w/in 1 year, mean ± SD** | 46.7±27.6 | 46.6±27.4 | 0.0036 |
| **CAN Score† for mortality w/in 1 year group (%)** |  |  |  |
| 0-30 | 33 | 32.9 | 0.0021 |
| 31-55 | 28.6 | 28.4 | 0.0044 |
| 56-75 | 20.2 | 20.2 | 0.0000 |
| 76-90 | 13.1 | 12.9 | 0.0059 |
| 95-96 | 1.6 | 1.5 | 0.0081 |
| 97-98 | 1.5 | 1.4 | 0.0084 |
| 99 | 0.6 | 0.5 | 0.0135 |
| Missing | 1.4 | 2.1 | -0.0534 |
| **Immunosuppressant medications\* (%)** | 5.3 | 5.2 | 0.0045 |

**Supplementary Figure 1.**
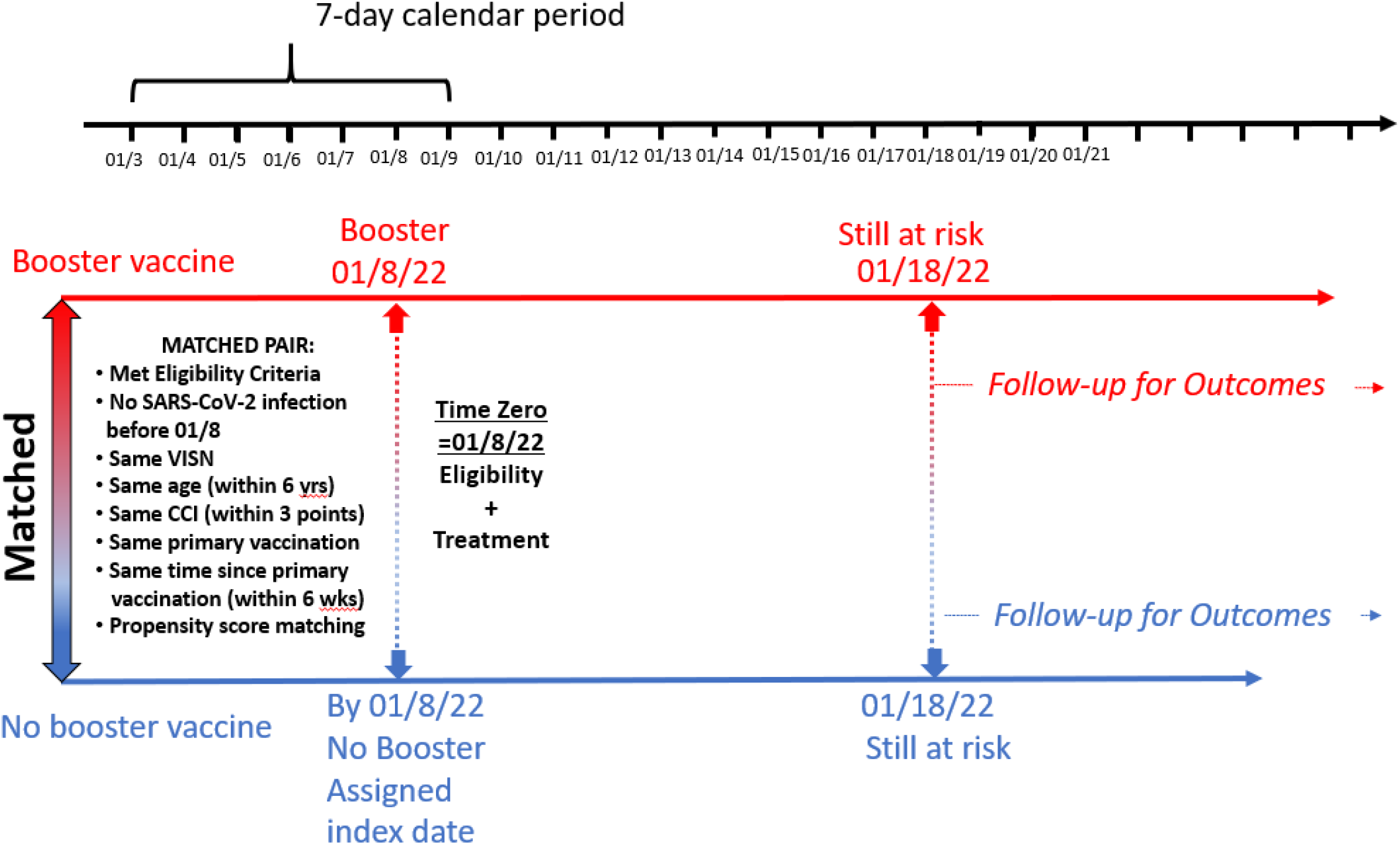
Schematic representation of the method used to match person(s) who did not receive a booster mRNA vaccine to each person who received a booster mRNA vaccine, within each 1-week period during the target trial period. The schematic shows a specific person during the 1-week period from 01/3/2022 to 01/9/2022 who received a booster mRNA vaccine dose on 01/8/1022 and met all of the target trial’s eligibility criteria and did not have SARS-CoV-2 infection as of 01/8/2022. This person is matched to a person who did not receive a booster vaccine dose as of 01/8/2022 (i.e. had only received two mRNA doses), who also fulfilled all the eligibility criteria and was alive as of 01/8/2022, and had the same VISN, type of primary vaccination, week of primary vaccination completion (within 6 weeks), age (within 6 years) and Charlson Comorbidity Index (CCI, within 3 points) as the booster dose recipient, and has a propensity score within 0.2 standard deviation of the mean (sdm) of that of the booster dose recipient. In practice, the propensity score difference between matched sets ranged from 0.0002-0.0033 sdm. Both persons in the matched pair have to remain at risk (i.e. alive, uninfected) as of the date 10 days later, after which outcomes are ascertained for the calculation of vaccine effectiveness

**Supplementary Figure 2.**
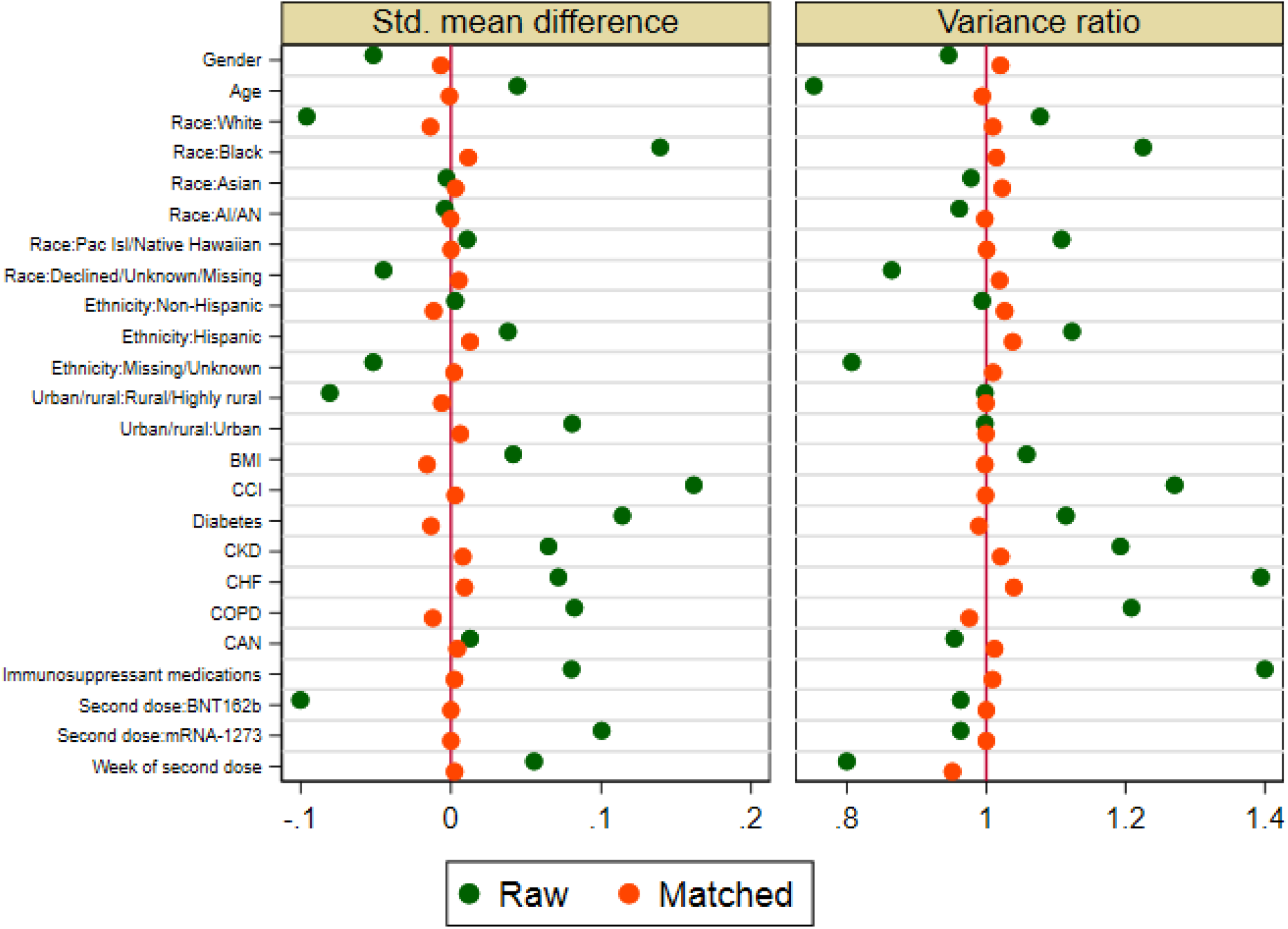
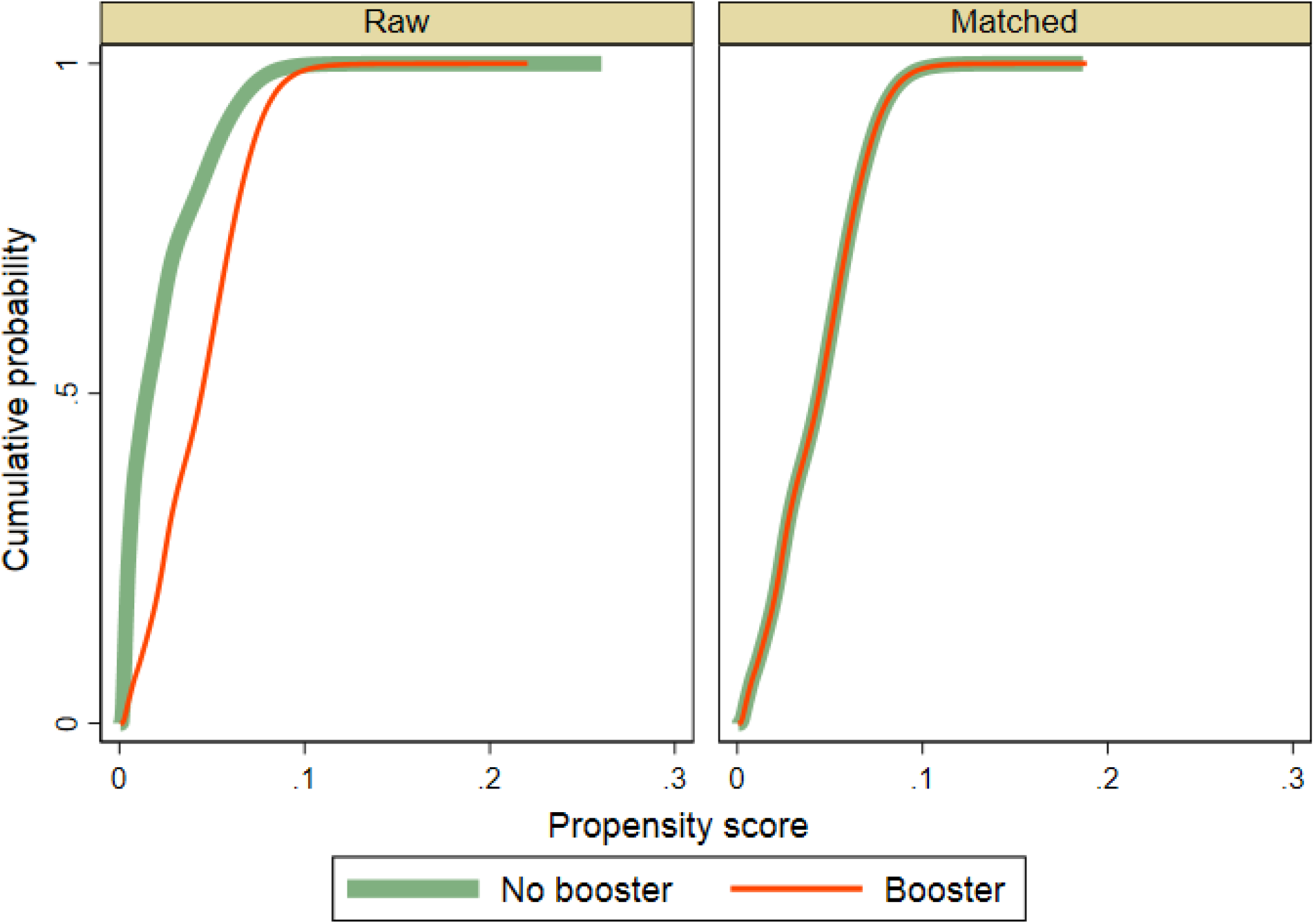
Comparison of baseline characteristics of persons who received a booster dose mRNA COVID-19 vaccine and their comparators who did not, before and after matching, demonstrate excellent balance after matching. **a.**Absolute standardized mean difference and variance ratio of baseline characteristics between booster dose mRNA vaccination recipients versus no booster dose shown for the raw (green dots) and matched (orange dots) data demonstrate balance after matching. Prior to matching, the absolute standardized difference in baseline characteristics between the 3^rd^ dose and no 3^rd^ dose groups ranged from 0.003-0.162 with a median of 0.060 (IQR: 0.040-0.089). After matching, the absolute standardized differences ranged from 0.000-0.016 with a median of 0.001 (IQR: 0.002-0.011). **b.** Cumulative distribution of propensity score between booster dose mRNA vaccination recipients versus no booster dose shown for the raw and matched data demonstrate balance after matching.

**Supplementary Figure 3.**
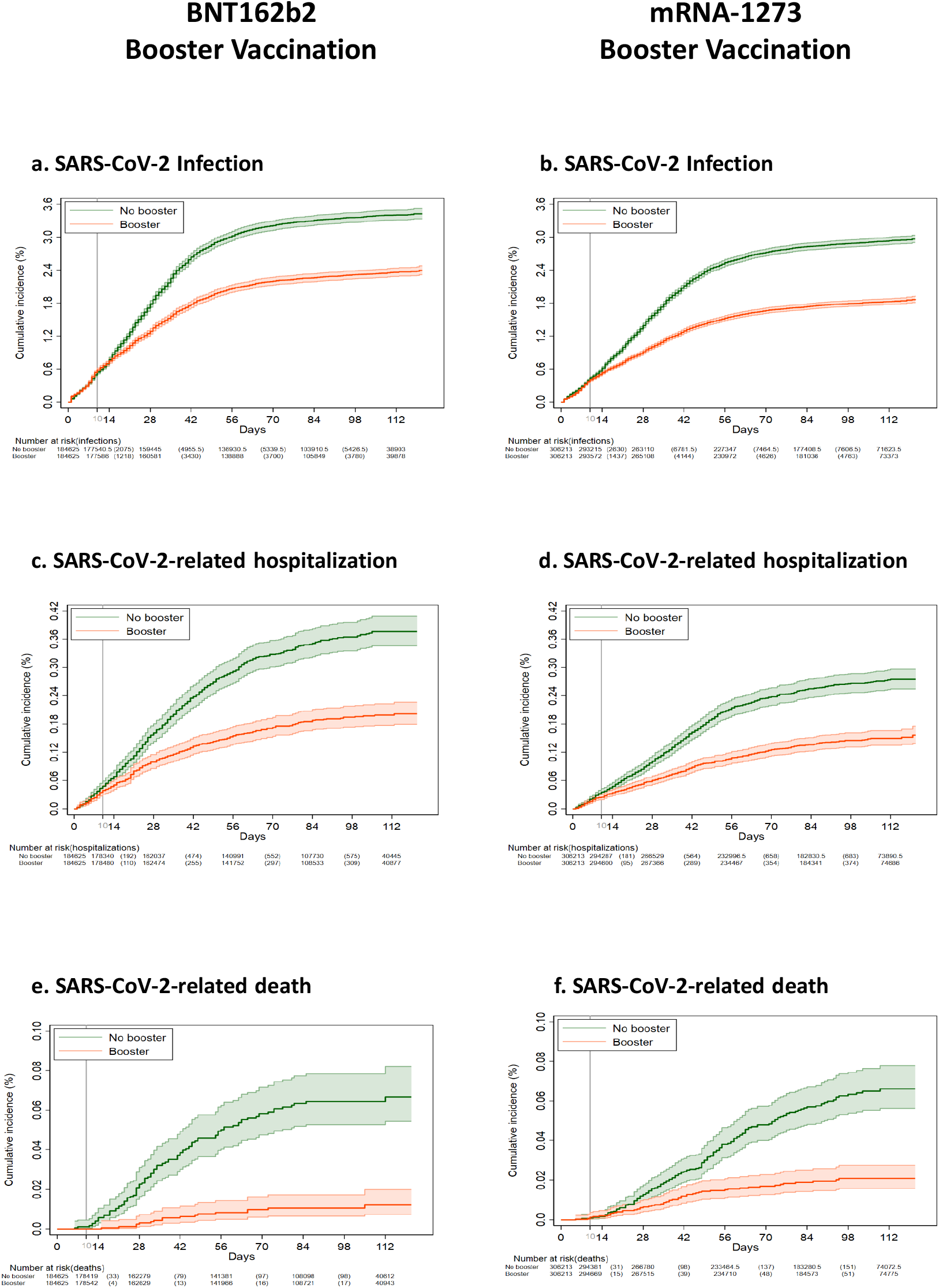
Kaplan-Meier curves comparing persons who received a booster mRNA COVID-19 vaccine versus their matched counterparts who did not with respect to the cumulative incidence (%) and 95% confidence intervals of SARS-CoV-2 infection (a) (b), SARS-CoV-2-related hospitalization (c)(d) and SARS-CoV-2-related death (e)(f). Results are shown separately for booster vaccination with BNT162b2 (a,c,e) and mRNA-1273(b,d,f)

